# Documented Human Implementation of NIH SBIR/STTR-Funded Neurosurgical Technologies: A Retrospective Cohort Study

**DOI:** 10.64898/2026.11.13.26368058

**Authors:** Abhinav Bachu, Raghav Rajesh, Shiau-Sing Ciecierska, Yoo Jin Ahn, Anand V. Germanwala

**Affiliations:** Department of Neurological Surgery, Loyola University Chicago Stritch School of Medicine, 2160 S. First Avenue, Maywood, IL 60153, United States

**Keywords:** Human implementation, Medical device translation, Neurosurgery, Research funding, Small Business Innovation Research, Small Business Technology Transfer

## Abstract

**Background:** Federal small business awards from the National Institutes of Health fund neurosurgical devices and procedural technologies, but how often the specific funded capability reaches human use is unknown. We determined how often these technologies had documented human implementation and in which settings.

**Methods:** We assembled a retrospective public-source cohort of technologies supported by Small Business Innovation Research and Small Business Technology Transfer awards from fiscal years 2005–2020 by keyword filtering and funded-purpose review. Confirmation required living-human use of the funded capability or a supported direct extension after a qualifying same-technology award, with public evidence by October 8, 2026. Exploratory comparisons examined program, observed phase, award era, and cumulative support. A 500-family random sample assessed screening omissions.

**Results:** The cohort comprised 209 technologies, 221 award families, and 453 applications receiving $206,825,343. Human implementation was documented for 31/209 technologies (14.8%; Wilson 95% confidence interval, 10.7%–20.3%); evidence was not located for 156 and was indeterminate for 22. Confirmed settings were neurosurgical-scope care (26), other clinical care (2), mixed surgical cohorts (1), and nonclinical human evaluation (2). Confirmation was more frequent with observed Phase II support than Phase I only (21/98 vs 10/111; difference, 12.4 percentage points; company-component bootstrap 95% confidence interval, 2.8–22.4). None of four conventional tests met a Holm-adjusted .05 threshold. Screening scenarios estimated 40–76 omitted families.

**Conclusion:** Approximately one in seven included technologies had documented human implementation. Linking individual awards to implemented capabilities provides an empirical benchmark for early neurosurgical translation and a practical foundation for tracking subsequent clinical evaluation.

## Introduction

Human implementation establishes that a capability developed in the laboratory has been exercised in a living person, allowing its clinical performance to be studied. The IDEAL and IDEAL-D frameworks locate early human evaluation within a staged process extending through comparative assessment and long-term surveillance.[7,16] Tracking human implementation shows which technological approaches have reached human evaluation and where further evidence is needed.

Research on translation has followed promising basic-science discoveries into clinical trials and licensed use.[3] Within neurosurgery, patent and publication analyses characterize innovation activity, while studies applying IDEAL assess the quality and sequence of evaluation.[4,9,15] The NIH Small Business Innovation Research (SBIR) and Small Business Technology Transfer (STTR) programs offer an additional starting point: awards identify a proposed capability, its intended purpose, and the organization developing it.[12] National assessments and linked-database evaluations have examined commercialization, patents, and organizational outcomes, [10,14] and recent neurosurgical research has mapped R01 funding among neurosurgeon-scientists.[13] Linking each funded capability to human use shows whether it reached human evaluation.

We used the technology as the unit of assessment. A company can hold several awards and develop related platforms with different mechanisms, delivery routes, and intended applications. Linking awards to the capability actually implemented makes it possible to follow a specific development pathway as products evolve. It also accommodates a funded neurosurgical capability that first reaches human evaluation in another clinical setting. Publication-defined surgical-device cohorts provide a precedent for tracing this transition,[5] while award-defined cohorts connect the transition to a documented research investment.

We evaluated neurosurgical technologies supported by NIH SBIR/STTR awards in fiscal years 2005–2020. Our primary objective was to estimate the proportion with qualifying public evidence of human implementation by October 8, 2026. We described the settings of confirmed implementation and explored associations with award characteristics. We also assessed potential screening omissions. We distinguished technologies without qualifying evidence from those with a specific unresolved question about the evidence.

## Materials and Methods

### Study design and cohort assembly

We conducted a retrospective cohort study using NIH RePORTER award records and public implementation evidence. We retrieved fiscal-year 2005–2020 R41/R42/R43/R44 records through the RePORTER API, partitioned by year, activity code, and 25 NIH institutes/centers.

[11] Acquisition yielded 29,658 applications representing 15,732 award families. We applied keyword filters to project titles, abstracts, and public-health-relevance text to identify candidates for funded-purpose review. Initial filtering flagged 3,336 families and filtered out 12,396; subsequent review and recovery established the study cohort. Supplementary Methods S1 lists the filter terms and acquisition settings. We used application identifiers, project numbers, recipient names, budget dates, and award amounts to link and characterize the records. Reporting followed applicable STROBE domains.[17]

Eligibility covered cranial or spinal intervention, neurovascular intervention, cerebrospinal fluid (CSF)/shunt or brain-directed neurocritical procedural management, neural repair interfaces, and implanted/percutaneous pain interfaces. Specifically funded adjuncts and manufacturing capabilities integral to these procedures were eligible. Funded purpose and procedural capability determined scope, including relevant peripheral nerve and pain procedures performed across specialties. Generic neurological benefit, systemic resuscitation, generic cell manufacturing, and disease modeling alone were insufficient. Supplementary Methods S1 details the scope rules.

We deduplicated applications by exact identifier and grouped them into technologies when the documentation supported a common mechanism and intended capability. Mixed-purpose families were allocated at application level. Eight families with unresolved eligibility were held outside the cohort. Figure 1 summarizes cohort assembly and screening dispositions. Figure 2 shows implementation classifications and settings. Figure S1 presents the separate exclusion-frame audit, which assesses omissions in the same 15,732-family universe after removing 378 previously covered families, leaving a 15,354-family audit frame. This frozen frame includes 21 families subsequently represented in the final cohort.

**Figure 1:**
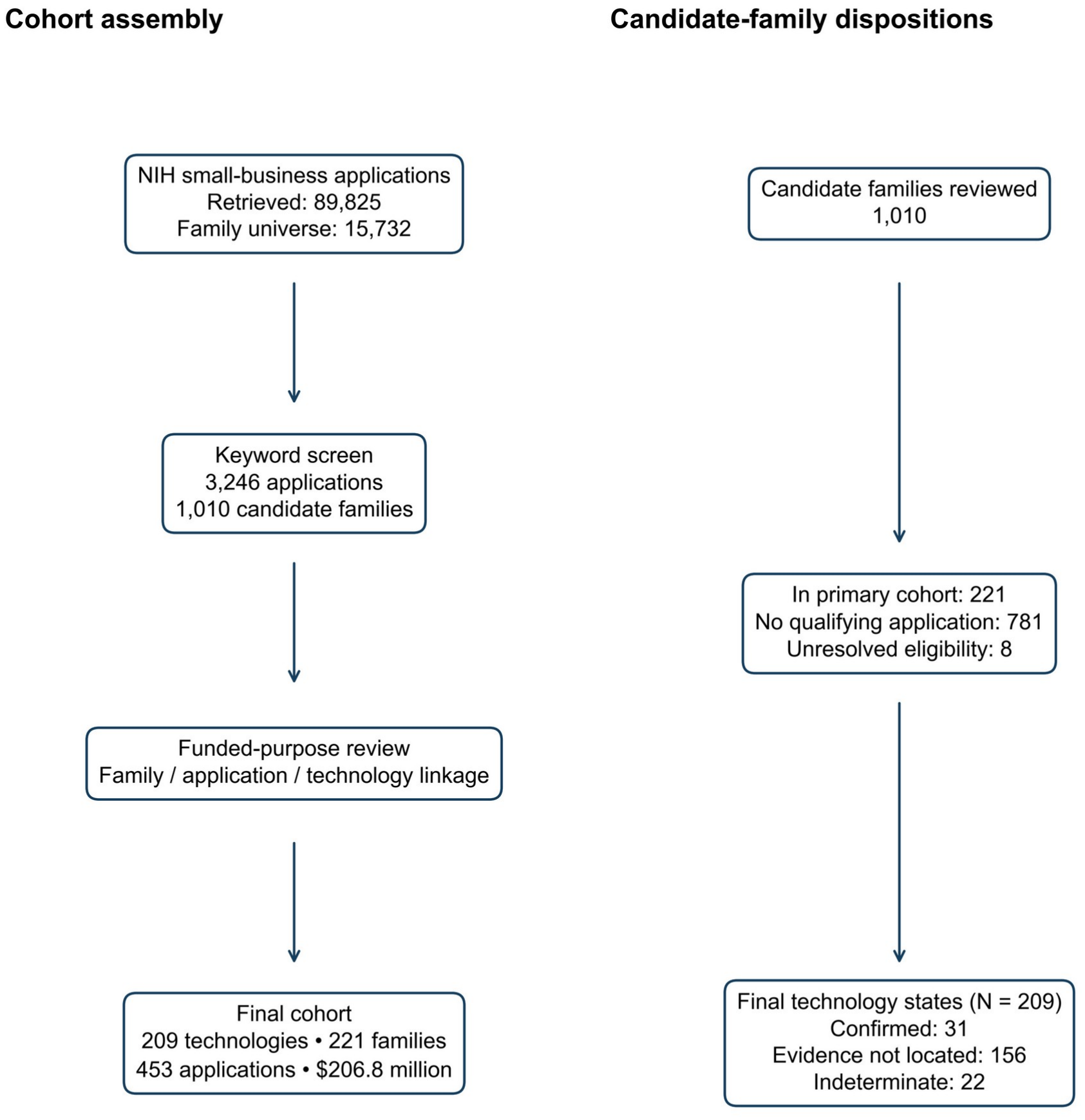
Cohort assembly and screening dispositions. Cohort assembly from NIH fiscal-year 2005–2020 award records. Current dispositions are shown within the original keyword-screening branches, allowing recovery from initially filtered families. The flagged branch includes 577 provisional candidates and 2,759 assessment candidates. Across both branches, 221 families were included, 8 remained on eligibility hold outside the cohort, 702 had source-linked documented exclusions, and 14,801 retained their historical screening exclusions. The latter retain the historical audited screening ledger’s ‘exclude’ status without a later family-specific source adjudication in the assembled review records; they are not independently source-verified exclusions. Source-linked documented exclusions combine retained historical and current family-specific decisions. Only included families converge into the cohort. Application-level allocation retained 453 applications, grouped into 209 technologies; these counting units are not interchangeable. NIH=National Institutes of Health; SBIR=Small Business Innovation Research; STTR=Small Business Technology Transfer; FY=fiscal year.

**Figure 2:**
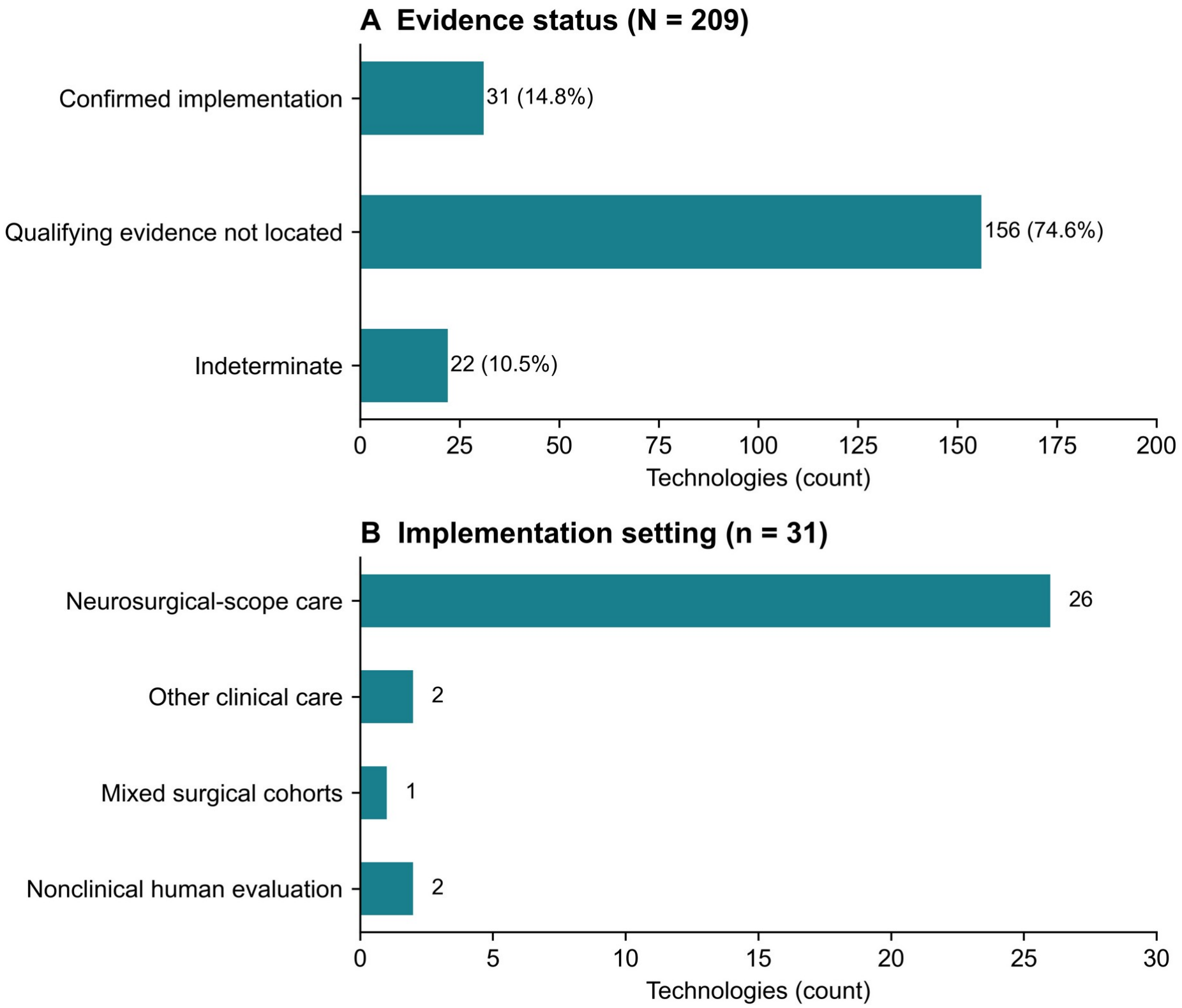
Evidence status and implementation setting in the 209-technology cohort. Panel A shows mutually exclusive implementation states among all 209 technologies; panel B shows mutually exclusive qualifying settings among the 31 confirmed technologies. Confirmation required actual living-human use of the funded capability or a supported direct extension after a qualifying same-technology award, with public evidence available by October 8, 2026. Qualifying evidence not located means no qualifying evidence and no concrete unresolved material lead after documented searches, not established nonuse. Indeterminate denotes a specific unresolved identity, chronology, or exposure fact. Confirmation was 31/209 (14.8%; Wilson 95% confidence interval, 10.7–20.3%); this interval assumes independent technologies. Neurosurgical-scope care includes relevant peripheral nerve and pain procedures across specialties. Other clinical care, mixed surgical cohorts, and nonclinical human evaluation identify distinct settings of qualifying use. Both panels use labeled count axes, with separate ranges of 0–200 technologies in A and 0–30 in B. Panel A gives counts and percentages; panel B gives counts only. Percentages may not sum to 100 because of rounding.

### Human implementation and evidence assessment

Confirmation required actual use of the funded capability or a supported direct extension in a living human after a qualifying same-technology award, with supporting public evidence available by October 8, 2026. Primary clinical reports, informative primary conference reports, and posted trial results could establish exposure. Regulatory authorization, sponsor claims, and registry status alone were insufficient. Animal, cadaver, specimen-only, and stored-data-only evaluations were excluded. Other-specialty clinical use and nonclinical human evaluation qualified when they exercised the funded capability.

We searched PubMed/Europe PMC, ClinicalTrials.gov, primary reports, and supporting public sources using grant identifiers, product names, supported aliases, company names, and mechanism-specific terms. We assessed capability identity, human exposure, and chronology together. Later retrievals, including October 14 verification searches, could establish that evidence was available by the October 8 cutoff. Search records retained source links and the facts supporting each decision.

We classified technologies as confirmed, not located, or indeterminate. Not located indicated no qualifying evidence and no concrete unresolved material lead after the documented searches. Indeterminate indicated a specific unresolved identity, chronology, or exposure fact that could change classification. All three groups remained in the primary denominator. We assigned confirmed technologies to four mutually exclusive settings: neurosurgical-scope care, other clinical care, mixed surgical cohorts, or nonclinical human evaluation.

One author performed evidence verification and review, with source identification and vetting assistance from a second author. Tables S5–S7 document confirming evidence, indeterminate endpoints, and eligibility holds.

### Award characteristics

We compared any STTR support, including both-program technologies, with SBIR only; any observed Phase II application (R42/R44) with Phase I only; and earliest included fiscal year 2005–2012 with 2013–2020. Cumulative support summed deduplicated qualifying application amounts in nominal US dollars. Observation opportunity extended from the earliest valid included budget start to the evidence cutoff. A separately verified earlier same-technology award could establish qualifying implementation chronology. Missing amounts and invalid or missing dates were not imputed.

### Statistical analysis

We calculated the confirmed proportion among all included technologies with a Wilson 95% confidence interval (CI).[18] Exploratory categorical comparisons used two-sided Fisher exact tests. Cumulative support was compared between confirmed technologies and the combined not-located/indeterminate group using a two-sided asymptotic Mann–Whitney test with tie and continuity correction. Holm adjustment covered the four comparisons. Funding effect sizes were the median difference and probability of superiority, defined as the probability that support for a confirmed technology exceeded support for an unconfirmed technology, with half credit for ties.

We estimated effect-size CIs with 10,000 whole company-component bootstrap replicates per contrast, resetting NumPy PCG64 seed 20261014 for each contrast. Technologies sharing normalized recipient identities were linked transitively, and all technologies in each sampled component were retained. CIs used linearly interpolated percentile limits. Wilson intervals and conventional tests assumed independent technologies; bootstrap intervals accounted for recorded shared recipients and were not multiplicity-adjusted. Analyses used Python, NumPy, and SciPy. Supplementary Methods S3 provides the calculation details.

#### Screening assessment and indeterminate scenario

We assessed potential omissions using a simple random sample without replacement of 500 families from a 15,354-family exclusion frame. Alternative classifications of unresolved families generated eligible-frame estimates and conditional exact hypergeometric limits. We incorporated deduplicated known recoveries and eligible families outside the cohort to estimate remaining omissions and family coverage. Supplementary Methods S4 describes the classification-sensitivity calculations. We also calculated a scenario in which all 22 indeterminate technologies became confirmed while the other classifications remained unchanged (Table S8).

### Ethics and AI assistance

Institutional review board review was not required at Loyola University Chicago Stritch School of Medicine for this public-source study. During the preparation of this work, the authors used ChatGPT (Codex, OpenAI; October 16–19, 2026) to assist with language editing and with writing analysis code. AI tools did not independently generate content; all text, code, and analyses were directed, reviewed, edited, and verified by the authors, who take full responsibility for the content of the publication.

## Results

### Cohort characteristics

The cohort comprised 209 technologies, 221 award families, and 453 applications receiving $206,825,343 in nominal support (Figure 1). There were 161 connected company components. Median observation opportunity was 14.3 years (range, 6.0–21.8). Funding amounts, valid budget starts, recipient information, and program, phase, and era classifications were complete. Sixteen budget starts fell outside the fiscal-year boundaries of their administrative records and were retained as recorded. One confirmed implementation preceded its earliest included budget start but followed a separately verified earlier same-technology award.

Spine surgery/reconstruction, neuro-oncology, and functional neurosurgery/epilepsy were the largest funded-purpose categories, with 32, 31, and 29 technologies, respectively (Table 1). There were 166 SBIR-only technologies, 39 STTR-only technologies, and 4 receiving both programs. Ninety-eight technologies had observed Phase II support and 111 had Phase I only. Table S4 provides the complete roster.

**Table 1:**
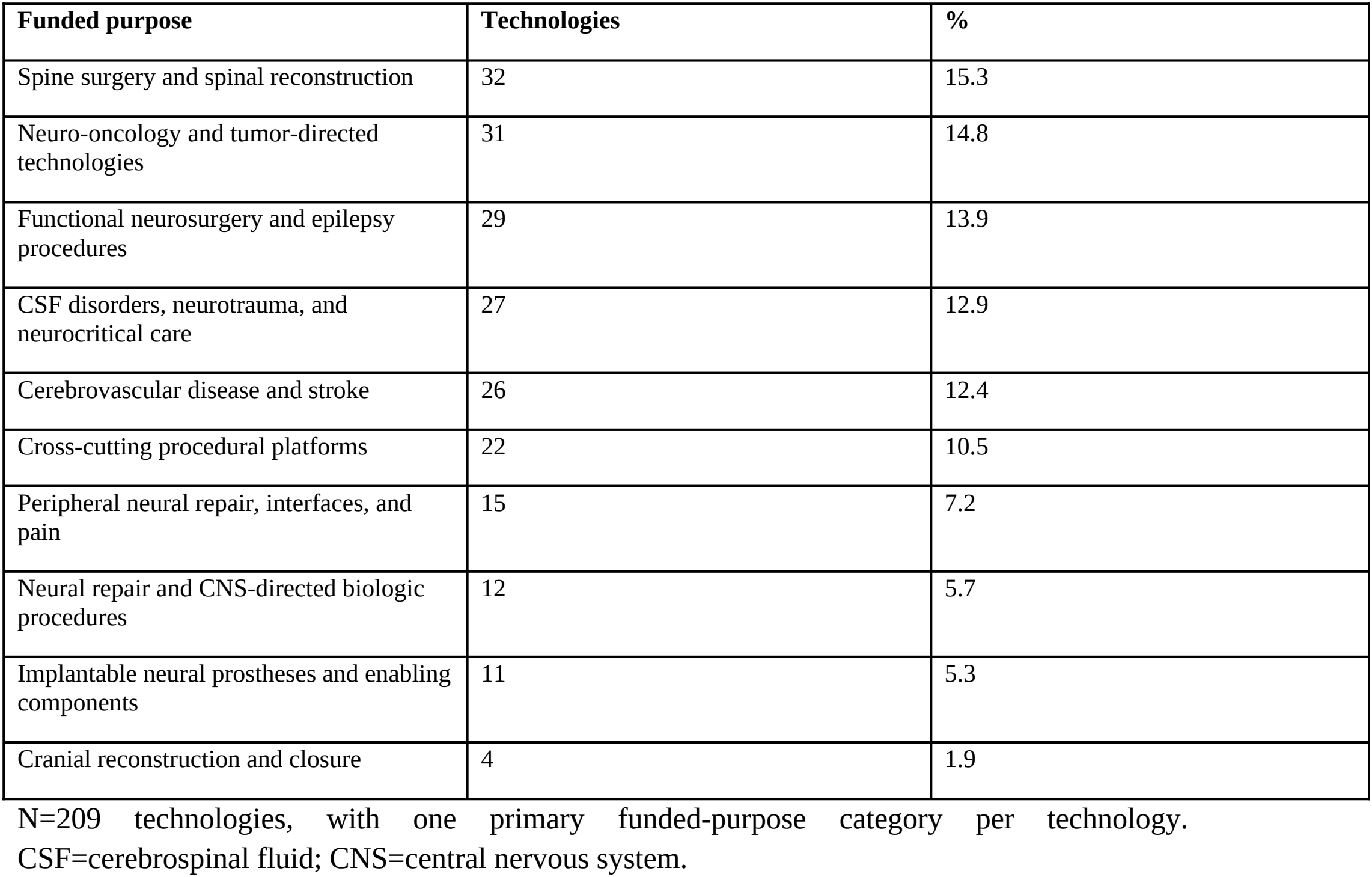
Primary funded-purpose categories.

| **Funded purpose** | **Technologies** | **%** |
|---|---|---|
| Spine surgery and spinal reconstruction | 32 | 15.3 |
| Neuro-oncology and tumor-directed technologies | 31 | 14.8 |
| Functional neurosurgery and epilepsy procedures | 29 | 13.9 |
| CSF disorders, neurotrauma, and neurocritical care | 27 | 12.9 |
| Cerebrovascular disease and stroke | 26 | 12.4 |
| Cross-cutting procedural platforms | 22 | 10.5 |
| Peripheral neural repair, interfaces, and pain | 15 | 7.2 |
| Neural repair and CNS-directed biologic procedures | 12 | 5.7 |
| Implantable neural prostheses and enabling components | 11 | 5.3 |
| Cranial reconstruction and closure | 4 | 1.9 |

N=209 technologies, with one primary funded-purpose category per technology.
CSF=cerebrospinal fluid; CNS=central nervous system.

#### Human implementation

Human implementation was documented for 31/209 technologies (14.8%; Wilson 95% CI, 10.7%–20.3%). Evidence was not located for 156/209 (74.6%) and was indeterminate for 22/209 (10.5%) (Table 2; Figure 2). Among confirmed technologies, 26 (83.9%) qualified through neurosurgical-scope care, 2 (6.5%) through other clinical care, 1 (3.2%) through a mixed surgical cohort, and 2 (6.5%) through nonclinical human evaluation. If all 22 indeterminate technologies became confirmed while the remaining classifications stayed unchanged, the proportion would be 53/209 (25.4%; Table S8).

**Table 2:** Human implementation and qualifying setting.

| **Ascertainment state or setting** | **n/N** | **%** |
|---|---|---|
| Confirmed | 31/209 | 14.8 |
| Not located | 156/209 | 74.6 |
| Indeterminate | 22/209 | 10.5 |
| Neurosurgical-scope care | 26/31 | 83.9 |
| Other clinical care | 2/31 | 6.5 |
| Mixed surgical cohort | 1/31 | 3.2 |
| Nonclinical human evaluation | 2/31 | 6.5 |

Ascertainment states use N=209; mutually exclusive implementation settings use N=31 confirmed technologies. The confirmed proportion has a Wilson 95% CI of 10.7%–20.3%. Settings follow procedural scope, including relevant peripheral nerve and pain care across specialties.

#### Exploratory award associations

Confirmation occurred in 7/43 technologies with any STTR support and 24/166 with SBIR only (16.3% vs 14.5%; difference, 1.8 percentage points; company-component bootstrap 95% CI, −10.0 to 15.2). Technologies with observed Phase II support had more confirmations than those with Phase I only (21/98 vs 10/111; 21.4% vs 9.0%; difference, 12.4 percentage points; 95% CI, 2.8–22.4) (Figure 3). The corresponding raw Fisher p value was .018 and Holm-adjusted p value was .073. Earlier-era technologies had confirmation in 21/111 compared with 10/98 later-era technologies (18.9% vs 10.2%; difference, 8.7 percentage points; 95% CI, −0.7 to 18.6). No comparison met a Holm-adjusted .05 threshold (Table 3; Figure 3).

**Table 3:** Exploratory categorical comparisons.

| **Contrast (first vs second)** | **Confirmed, n/N (%)** | **Difference, pp (bootstrap 95% CI)** | **Raw p** | **Holm p** |
|---|---|---|---|---|
| Program<br>Any STTR vs SBIR only | 7/43 (16.3%)<br>vs 24/166 (14.5%) | 1.8<br>(-10.0 to 15.2) | 0.810 | 0.810 |
| Observed phase<br>Any Phase II vs Phase I only | 21/98 (21.4%)<br>vs 10/111 (9.0%) | 12.4<br>(2.8 to 22.4) | 0.018 | 0.073 |
| Earliest included fiscal year<br>2005–2012 vs 2013–2020 | 21/111 (18.9%)<br>vs 10/98 (10.2%) | 8.7<br>(-0.7 to 18.6) | 0.083 | 0.249 |

Differences are first minus second group. Fisher tests assume independent technologies. Difference intervals resample whole company components and are not multiplicity-adjusted. Holm adjustment covers all four tests, including funding. Group-specific Wilson intervals and separate not-located/indeterminate counts appear in Table S3. STTR=Small Business Technology Transfer; SBIR=Small Business Innovation Research; pp=percentage points; CI=confidence interval.

**Figure 3:**
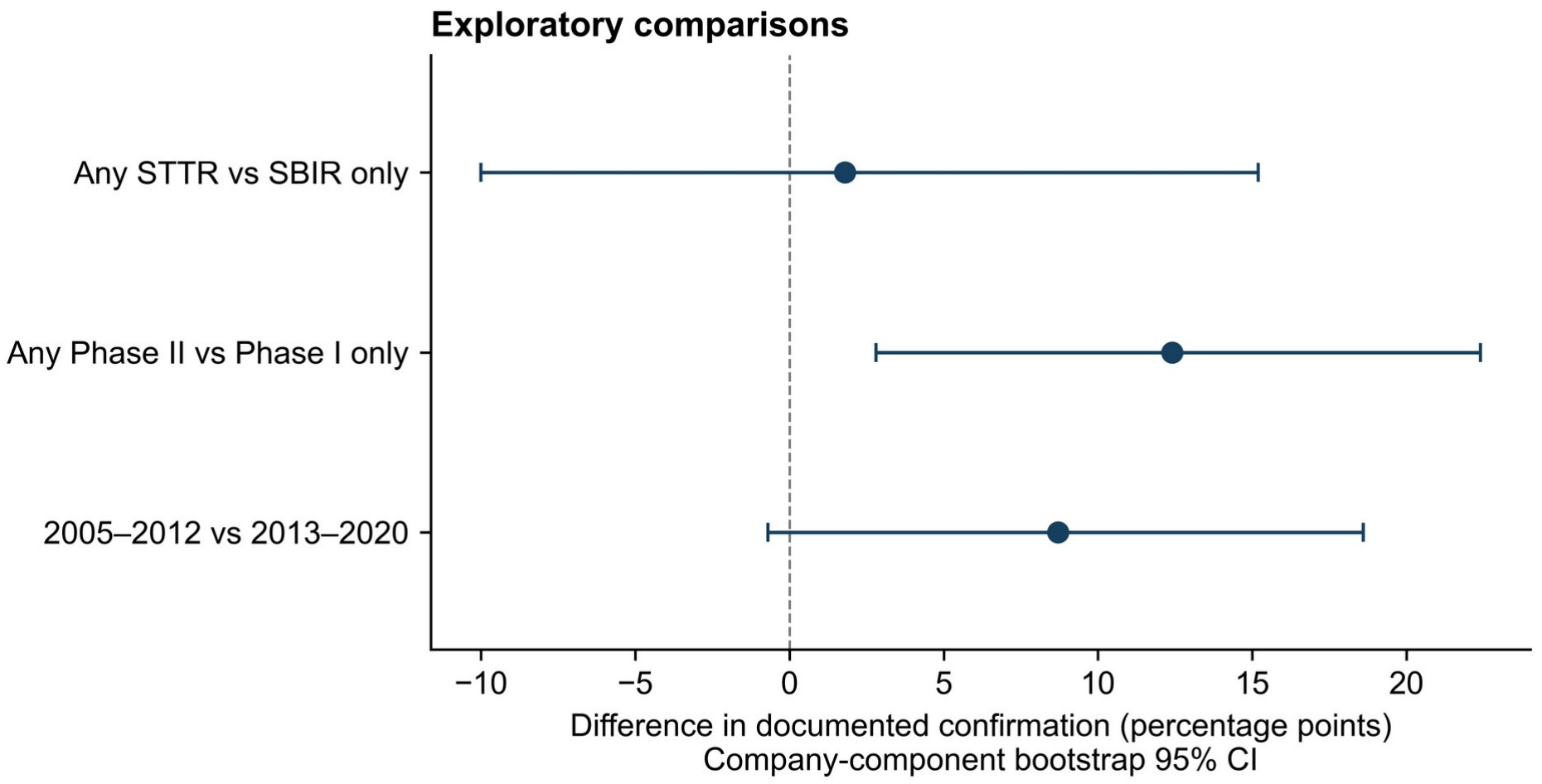
Exploratory differences in documented confirmation by program, phase, and award era. Dots show differences in documented confirmation between the first and second groups, in percentage points; horizontal lines show company-component bootstrap 95% confidence intervals. Counts are confirmed technologies/group denominator; qualifying-evidence-not-located and indeterminate technologies remain in each denominator. Any STTR includes STTR-only and both-program technologies; any observed Phase II means at least one included R42/R44 application. Award era is earliest included fiscal year, 2005–2012 versus 2013–2020. Intervals use 10,000 whole-component percentile bootstrap replicates from 161 recorded-recipient components and are not multiplicity-adjusted; software and random-seed details are provided in Supplementary Methods S3. Conventional Fisher tests assume independent technologies; Holm adjustment covers program, phase, era, and funding. No conventional test met the adjusted .05 threshold; the Phase II adjusted p value is .073 despite its bootstrap interval excluding zero. The dashed line marks zero difference. These associations are exploratory, not causal. SBIR=Small Business Innovation Research; STTR=Small Business Technology Transfer; CI=confidence interval.

Median cumulative support was $1,124,596 for confirmed technologies and $400,053 for the 178 without documented confirmation (Table 4). The median difference was $724,543 (company-component bootstrap 95% CI, −$131,044 to $1,458,154), and probability of superiority was 0.588 (95% CI, 0.458–0.712). The Mann–Whitney U statistic was 3,244, with raw p=.119 and Holm-adjusted p=.249.

**Table 4:** Cumulative nominal support by ascertainment state.

| **Group** | **N** | **Mean (SD)** | **Median (Q1–Q3)** | **Range** |
|---|---|---|---|---|
| All technologies | 209 | 990 (1,079) | 431 (223–1,617) | 77–5,169 |
| Confirmed | 31 | 1,405 (1,312) | 1,125 (211–2,014) | 100–4,724 |
| Not located | 156 | 920 (1,043) | 383 (224–1,431) | 77–5,169 |
| Indeterminate | 22 | 899 (864) | 509 (228–1,485) | 118–3,333 |
| No documented confirmation | 178 | 917 (1,020) | 400 (224–1,468) | 77–5,169 |

Amounts are thousands of nominal US dollars, rounded to the nearest thousand. All values are known: missing=0 in every row. SD uses n−1 and quartiles use linear interpolation. No documented confirmation combines not located and indeterminate. The funding test gives U=3,244; raw p=.119; Holm p=.249. Median difference=\$724,543 (company-component bootstrap 95% CI, −\$131,044 to \$1,458,154); probability of superiority=0.588 (95% CI, 0.458–0.712).

#### Screening assessment

The 500-family sample contained 2 eligible, 497 excluded, and 1 unresolved family. Under unresolved-excluded and unresolved-eligible scenarios, estimated remaining omissions were 40 and 76 families, corresponding to family coverage of 84.5% and 74.4%. The combined transformed classification-sensitivity envelope was 0–248 remaining families and 47.1%–100.0% coverage (Table S2).

## Discussion

Human implementation was documented for approximately one in seven included NIH-funded neurosurgical technologies. By linking individual awards to implemented capabilities, this study provides an empirical account of which technological approaches reached human evaluation and in what settings. Each award-to-capability link connects a funding record to a specific human-use report, allowing subsequent clinical evidence for that capability to be followed. The not-located group requires further searches for qualifying evidence. The indeterminate group has specific unresolved facts that could change classification.

This award-defined approach complements earlier translation studies. Contopoulos-Ioannidis and colleagues followed promising basic-science findings toward clinical trials and licensed use,[3] while Marcus and colleagues identified first-in-human reports for 24 of 205 publication-defined surgical devices (11.7%) and estimated a 9.8% probability of translation at 10 years.[5] Our cohort begins with NIH-supported capabilities and measures documented implementation through a common cutoff. Starting from NIH awards also includes technologies without located human-use publications. Linking awards and human-use reports adds a funding-based perspective to studies organized around publications, patents, or company outcomes.

The human-use reports illustrate why capability-level linkage matters. FLoupe imaging was exercised during glioma operations,[8] and silicon-nitride spacers were implanted in the CASCADE cervical-fusion trial.[1] Multimodal scanning-fiber endoscopy was evaluated in 22 participants with Barrett’s esophagus, demonstrating a clinical extension outside neurosurgical care.[2] Following the mechanism, configuration, and route of use allows a technology’s development to remain traceable across changing product names and clinical indications.

The next evaluative question is the quality of the clinical evidence generated after implementation. Neurosurgical IDEAL reviews have shown gaps in staged evaluation: Muskens and colleagues examined the introduction of endoscopic endonasal meningioma surgery and WEB devices,[9] and Ota and colleagues found appropriate IDEAL application in only 5 of 14 primary neurosurgical papers identified in their citation review.[15] Our roster offers a complementary starting population for following funded technologies into feasibility studies, comparative trials, and longer-term assessment. Retaining the distinction between clinical and nonclinical human evaluation makes that progression interpretable.

Future longitudinal studies could examine why confirmation was more frequent among technologies with observed Phase II support. As technologies mature, they may receive later-phase support and develop clinical partnerships. Additional documentation may also become available. Phase II awards and cumulative funding may also occur after implementation. A prospective study recording development stage and funding before human use could distinguish these pathways. Here, the bootstrap interval and Holm-adjusted test summarize different inferential procedures, and the phase association remains exploratory. Program and funding comparisons could also inform studies of development support and the organizational outcomes measured in prior NIH SBIR/STTR evaluations.[10,14]

Consistent technology identifiers could improve linkage across award records and human-use reports. Award records could carry stable product or prototype identifiers, links to earlier development awards, and dated references to human-use reports. Investigators could specify which funded capability was exercised and the procedure in which it was used. Trial and publication records could retain those identifiers as devices change ownership or name. These records would make it easier to follow development across specialties and assess whether the available evidence supports further clinical study. Human implementation, regulatory authorization, comparative benefit, and adoption would then be trackable as distinct milestones, consistent with IDEAL and IDEAL-D.[6,7,16]

### Limitations

Public-source ascertainment may miss unpublished or inaccessible use, and search coverage differed among technologies. Incorrect capability linkage or chronology could also affect classification. Some timing decisions relied on supported development sequences. Twenty-two endpoints remained indeterminate, and screening estimates indicated potential omissions with substantial uncertainty. The 14.8% therefore describes documented implementation in the assembled cohort rather than the full NIH portfolio. Observation opportunity varied, and included awards were not necessarily lifetime-first support. The broad endpoint encompasses other-specialty and nonclinical human use. Award comparisons were exploratory and temporally ambiguous, and company components captured recorded shared recipients rather than every corporate relationship. These features limit causal and portfolio-wide inference.

## Conclusion

Linking NIH SBIR/STTR awards to technology-specific human evidence identified implementation for 31 of 209 included neurosurgical technologies. This award-to-capability approach provides a benchmark for early translation and a traceable foundation for evaluating how funded technologies progress through subsequent clinical study.

## Data Availability

Study-generated data and code, NIH source records, screening and recovery ledgers, and dated search records are available from the corresponding author on reasonable request. Access to third-party materials is subject to applicable licenses and permissions. Where redistribution is restricted, we will provide citations, persistent identifiers, and source links. Supplementary Methods S5 describes the archived analytical materials. The data analyzed are public NIH RePORTER award records and published literature.

https://reporter.nih.gov

## Disclosures and availability

### Funding and competing interests

This study received no funding or sponsorship. The authors declare no competing interests.

### Data and code availability

Study-generated data and code, NIH source records, screening and recovery ledgers, and dated search records are available from the corresponding author on reasonable request. Access to third-party materials is subject to applicable licenses and permissions. Where redistribution is restricted, we will provide citations, persistent identifiers, and source links. Supplementary Methods S5 describes the archived analytical materials.

## Notes

### Competing Interest Statement

The authors declare no competing interests. The authors and their institutions have not received any payments or services in the past 36 months from a third party that could be perceived to influence, or give the appearance of potentially influencing, this work.

### Author Declarations

The source data were openly available to the public before the initiation of this study. NIH RePORTER award records are available at https://reporter.nih.gov; additional evidence was drawn from published literature.

