## Supplementary Material for "Documented Human Implementation of NIH SBIR/STTR-Funded Neurosurgical Technologies: A Retrospective Cohort Study"

**Supplementary Methods S1. Eligibility and technology linkage**

We retrieved NIH RePORTER records for fiscal years 2005–2020 and R41/R42/R43/R44 activity codes using POST requests to the v2 projects/search endpoint. Requests were partitioned by fiscal year, activity code, and 25 administrative NIH institutes/centers, with is_agency_admin=true, a page limit of 500, offset 0, and fiscal-year ascending order. The 1,600 completed request partitions yielded 29,658 applications representing 15,732 families, with no duplicate or conflicting application identifiers. Keyword filters were applied to the retrieved award text to identify candidates for funded-purpose review.

*Keyword filters*

We applied case-insensitive patterns to project titles, abstracts, and public-health-relevance text across award years. The domain term groups were:

Cranial: neurosurg, cranial, craniotom, brain tumor/tumour, brain surgery, brain resection, skull base, stereotact, and intracranial.

Spine: spinal/spine fusion, surgery, implant, instrumentation, decompression, or fixation; vertebr, intervertebral, laminectom, and pedicle screw.

Peripheral nerve: peripheral nerve; nerve repair, graft, conduit, transfer, decompression, or surgery; carpal tunnel; and brachial plexus.

Implanted pain interfaces: spinal cord stimulation, dorsal root ganglion stimulation, peripheral nerve stimulation, implanted neuromodulation, and intrathecal pump.

CSF/ICP: cerebrospinal fluid, CSF, hydrocephal, intracranial pressure, external ventricular drain, ventricular shunt, and CSF shunt.

Neurovascular: cerebral/intracranial aneurysm, neurovascular, cerebral arteriovenous malformation/AVM, mechanical thrombectomy, and intracranial stent, catheter, or embolization.

Functional/epilepsy: deep brain stimulation, epilepsy surgery, seizure focus/onset localization, responsive neurostimulation, functional neurosurgery, and stereo-EEG.

Oncology/skull base: brain tumor/tumour, glioma, glioblastoma, skull base, intracranial tumor/tumour, and pituitary tumor/adenoma.

Pediatric craniofacial: craniosynostosis, craniofacial surgery/reconstruction, pediatric neurosurgery, neural tube defect, and spina bifida.

Neurotrauma: traumatic brain injury, brain trauma, spinal cord injury, and cranial trauma.

Patterns accommodated word stems, spelling variants, and specified compound phrases. Procedural terms included surgery/surgical, operative, intraoperative, resection, ablation, implant, catheter, shunt, drainage, embolization, thrombectomy, decompression, fusion, fixation, instrumentation, biomaterial, graft, conduit, prosthesis, device, stimulation, neuromodulation, navigation, robot, image guidance, surgical/presurgical planning, trajectory, endoscopy, needle, clip, coil, stent, monitoring, pressure sensor, and flow sensor. Intended-use terms included use during/for treatment or management, intended for/to, clinical application/use/trial/evaluation, patient, procedure, therapy, treatment, and operation. A qualifying domain, procedure, and intended-use context in the same application generated a provisional candidate; ambiguous context was retained for further assessment. Systemic-drug, general diagnostic/rehabilitation, and research-only contexts informed filtering when a qualifying procedural purpose was absent. The complete regular expressions and combination rules are retained with the source code.

Initial keyword filtering produced 577 provisional candidates and 2,759 families requiring further funded-purpose assessment (3,336 combined), while 12,396 families were filtered out. These are initial filter dispositions. Funded-purpose review, recovery, and application-level allocation established the final cohort of 221 families, 453 applications, and 209 technologies. Eligibility required a substantive purpose involving cranial or spinal intervention, neurovascular intervention, CSF/shunt or brain-directed neurocritical procedural management, neural repair interfaces, or implanted/percutaneous pain interfaces. Specifically funded procedural adjuncts and manufacturing capabilities integral to those procedures could qualify before clinical readiness.

Generic neurological benefit, systemic resuscitation, generic cell manufacturing, and disease modeling alone were insufficient. Autonomic urology and cardiovascular technologies needed a separate qualifying neurosurgical purpose. General neuraxial anesthesia guidance, nonsurgical epidural analgesia, and entirely intraocular retinal prostheses were excluded. Technology categories describe primary funded purpose, rather than diagnoses, billing codes, clinical validation, or exclusive lifetime use.

An application was linked by its exact application identifier and allocated to one technology. Grouping required support for the same mechanism and intended capability. A common recipient or similar title did not independently establish identity. Mixed-purpose families were allocated at application level so that systemic-only applications did not qualify solely through another application in the family. Applications were deduplicated before technology-level aggregation. The included cohort contains 209 technologies, 221 families, and 453 applications. Eight unresolved families remain outside the denominator and are documented in Table S7. The audit frame was frozen as the 15,732-family acquisition universe minus 378 previously covered families, yielding 15,354 families (Figure S1; Supplementary Methods S4). Of the 221 families in the final cohort, 200 belonged to the covered set and 21 to the frozen audit frame.

Human confirmation required identity of the funded capability or a supported direct extension, actual living-human exercise of that capability, implementation after a qualifying same-technology award, and qualifying public evidence available by October 8, 2026. Primary clinical reports, informative primary conference reports, and posted trial results could establish exposure. Registry status, regulatory authorization, and sponsor claims alone could not. Animal, cadaver, specimen-only, and stored-data-only studies were outside the endpoint. Other clinical indications and nonclinical human evaluation could qualify if actual implementation of the funded capability was established.

The earliest included budget start defines observation opportunity. A separately verified earlier qualifying award can establish chronology even if implementation precedes the included start. One technology had that pattern. The other 208 pre-included-start flags were unknown, rather than verified negatives. Neither included fiscal year nor publication year was substituted for an exact implementation date. Supported development/use sequences were retained when exact dates were unavailable.

**Supplementary Methods S2. Public-source searches and evidence states**

Endpoint searches used grant identifiers, product names, supported aliases, company names as leads, and mechanism- or route-specific terms. Sources included PubMed, Europe PMC, ClinicalTrials.gov, primary clinical publications, informative conference reports, and supporting regulatory and public records. Public entry points are https://reporter.nih.gov/, https://pubmed.ncbi.nlm.nih.gov/, https://europepmc.org/, and https://clinicaltrials.gov/.

We used grant identifiers, product names and supported aliases to identify candidate human-use reports. Company names and mechanism- or route-specific terms supported additional searches and disambiguation. For each potential confirmation, we evaluated the funded capability, its relationship to the implemented technology, actual human exposure, and chronology relative to a qualifying award. Primary source quotations, links, dated retrievals, and unresolved facts were retained in the study archive.

The exposure and public-evidence cutoff was October 8, 2026. Subsequent retrievals, including October 14 verification searches, were used to assess evidence available by that cutoff. Search coverage and retrieval outcomes are documented in the dated records, including incomplete or failed retrievals.

One author performed evidence verification and review, with source identification and vetting assistance from a second author. Tables S5 and S6 document the grounds for confirmed and indeterminate classifications, respectively. Qualifying settings describe the procedural scope of implementation, including peripheral nerve and pain care delivered across specialties.

**Supplementary Methods S3. Variables, dependence, and statistical calculations**

Application-level funding was summed within technologies after deduplication. Total nominal support is selection-window support in US dollars without inflation adjustment. A missing application amount would make a technology total missing, with known subtotal retained separately; all 453 amounts were present in this cohort. Means, medians, standard deviations, and quartiles are technology-level summaries. Standard deviations use $n-1$ and quartiles use linear interpolation.

Program is SBIR only, STTR only, or both. Any STTR combines STTR-only and both-program technologies. Observed Phase II means at least one included R42 or R44 application; Phase I only means no included Phase II application. These definitions do not establish lifetime progression. Award era uses the minimum included fiscal year, split at 2012/2013. Observation opportunity in years is the number of days from the earliest valid included budget start to October 8, 2026, divided by 365.25. Missing, invalid, partial, post-cutoff, or start-after-end dates were not imputed. Fiscal-year discrepancies were flagged while retaining source budget dates.

Recipient identities were normalized using Unicode NFKC, uppercase conversion, punctuation, and whitespace rules while preserving legal suffixes and distinct words. No fuzzy matching was implied. Technologies sharing an identity were connected transitively into company components. This accounts for recorded shared recipients, rather than all possible corporate ownership, licensing, or acquisition relationships.

For k confirmations among n technologies, $p=k/n$. With $z=1.959963984540054$, the Wilson interval is centered at $c=(p+z^2/[2n])/(1+z^2/n)$, with half-width $h=z\sqrt{(p[1-p]/n+z^2/[4n^2])}/(1+z^2/n)$; its endpoints are $c-h$ and $c+h$. The primary calculation used $k=31$ and $n=209$. The interval conditions on the included cohort and observed classifications and assumes independent technologies.

Categorical effect sizes are $100(pA-pB)$ percentage points. Two-sided Fisher tests use confirmed and no-documented-confirmation counts in each group. The latter combines not located and indeterminate. The funding test is two-sided asymptotic Mann–Whitney with tie correction and continuity correction. Its U statistic is not a test solely of the median difference. Probability of superiority is $U/(nA \times nB)$, using the confirmed group as A and half credit for ties. The separately reported median difference is median(A)−median(B).

Holm adjustment covers program, phase, era, and funding. Order the four raw p values from smallest to largest. Multiply the ordered values by 4, 3, 2, and 1, respectively, apply a cumulative maximum, cap at 1, and restore the original order. This controls multiplicity within the conventional test family under its assumptions; it is not adjustment for confounding or correction of source ascertainment.

For each bootstrap contrast, component identifiers and technology identifiers were sorted. NumPy PCG64 was initialized with seed 20261014 separately for each contrast. Each replicate sampled 161 company components uniformly with replacement and retained every technology in each sampled component, including repeated copies when a component was sampled more than once. The 2.5th and 97.5th linearly interpolated percentiles of 10,000 valid replicates defined the intervals. Bootstrap contrasts required at least 20 components, at least 5 contributing to each group, and complete recipient identity. There were no empty-group replicates.

The bootstrap intervals address dependence represented in the recipient components and are not multiplicity-adjusted. They are not inverted Fisher tests. In particular, the phase difference's interval and Holm-adjusted p value do not provide interchangeable significance decisions. No regression, propensity-score model, causal weighting, or survival model was used.

**Supplementary Methods S4. Screening omission accounting**

A simple random sample without replacement of 500 families was drawn from the frozen 15,354-family exclusion frame, constructed as 15,732 retrieved families minus 378 previously covered families. Sampling used random.Random(20261014).sample(sorted(frame), 500). The sample contained 2 eligible families, 497 excluded families, and 1 unresolved family. The unresolved-excluded scenario uses $x=2$; the unresolved-eligible scenario uses $x=3$. Under a hypergeometric model with unknown eligible frame total M, the point estimate is $15{,}354 \times x/500$. Conditional two-sided exact limits are obtained by retaining M values for which both relevant tail probabilities are at least .025.

Known eligible and excluded sets constrain feasible frame totals under each scenario. Deduplicated eligible families already recovered from the frame must be subtracted once from the eligible-total estimate. Known or scenario-assumed eligible families absent from the cohort are then added to remaining unidentified families. Family coverage is 221/(221+remaining omitted families). Targeted retrievals are not a probability sample and were not extrapolated as if randomly sampled.

The scenario point estimates are 61 versus 92 eligible frame families, with raw conditional exact limits of 8–218 versus 20–264. These frame totals already include known recoveries. We incorporated 21 versus 24 known eligible frame families and 0 versus 8 known/scenario-assumed eligible families outside the cohort, yielding approximately 40 versus 76 remaining omissions. The combined transformed classification-sensitivity envelope is 0–248 remaining families and 47.1%–100.0% coverage. The archived feasible-set ledger defines the scenario-specific constraints. These transformed bounds describe classification sensitivity at the family level.

**Supplementary Methods S5. Completeness, diagnostic checks, and available data**

Funding amounts and valid budget starts were available for all 453 applications. Recipient information and program, phase, and era classifications were complete for all 209 technologies. We retained 16 budget starts falling outside the administrative fiscal-year boundaries. Median observation opportunity was 14.3 years (range, 6.0–21.8), and 12 technologies had more than one recorded recipient.

The 161 company components comprised 133 singletons, 17 components of size two, 7 of size three, 2 of size four, 1 of size five, and 1 of size eight. Components contributing to first/second groups numbered 40/124 for program, 77/101 for phase, 83/86 for era, and 29/139 for funding. A component could contribute to both groups. All four contrasts had 10,000 valid bootstrap replicates and zero empty-group replicates.

The archived analytical data include the technology roster, implementation evidence, unresolved endpoint facts, application covariates, technology covariates, and category definitions. Some CSV cells contain JSON-encoded lists. Blank chronology flags represent unknown values. The category dictionary includes one unused category, M; no included technology is assigned to it. Study IDs remain stable across cohort development, while the row numbers in Table S4 enumerate the 209 included technologies.

We verified the designated statistics from the packaged analytical inputs, including cross-table counts, funding aggregation, recipient components, observation opportunity, categorical tests, Holm adjustment, and bootstrap intervals. Analysis scripts, dependency records, and environment versions are documented with the archived code.

*Indeterminate classification scenario*

We examined the arithmetic consequence of confirming all 22 indeterminate technologies while retaining the 156 not-located classifications. The observed proportion was 31/209 (14.8%); the hypothetical proportion was 53/209

(25.4%). Table S8 reports this scenario. It is conditional on the stated reclassification and does not bound actual human implementation among technologies without located evidence.

*Data and code access*

The original study archive contains NIH award records, source-linked extraction and analysis code, dependency records, dated searches and retrievals, the original screening sample, and cohort/recovery ledgers. A separate reproducibility archive contains selected analytical inputs, source packets, search logs, numerical results, and verification records. The full source-to-result workflow uses the original acquisition, extraction, and dependency records.

Study-generated materials and source-access records are available from the corresponding author on reasonable request. Redistribution of third-party full text is governed by its license or permissions; where restricted, citations, persistent identifiers, and source links will be provided.

**Supplementary figure**

**Figure S1: Probability-sample screening audit and final cohort membership**

Panel A shows the screening-audit design. Removing 378 previously covered families from the 15,732-family acquisition universe defined a frozen 15,354-family frame. A simple random sample of 500 families without replacement (Python random.Random seed 20261014) contained 2 eligible, 497 excluded, and 1 unresolved family. Panel B shows overlap with final cohort assembly: 200 included families belonged to the covered set and 21 to the frozen frame, totaling 221. These are subsets, not additive audit yields. The frame and probability-sample denominator remain unchanged after recovery. Nonrandom targeted searches can overlap the sample and are deduplicated in cohort accounting, not pooled with the probability sample. Omission scenarios and classification-sensitive uncertainty appear in Table S2 and Supplementary Methods S4.

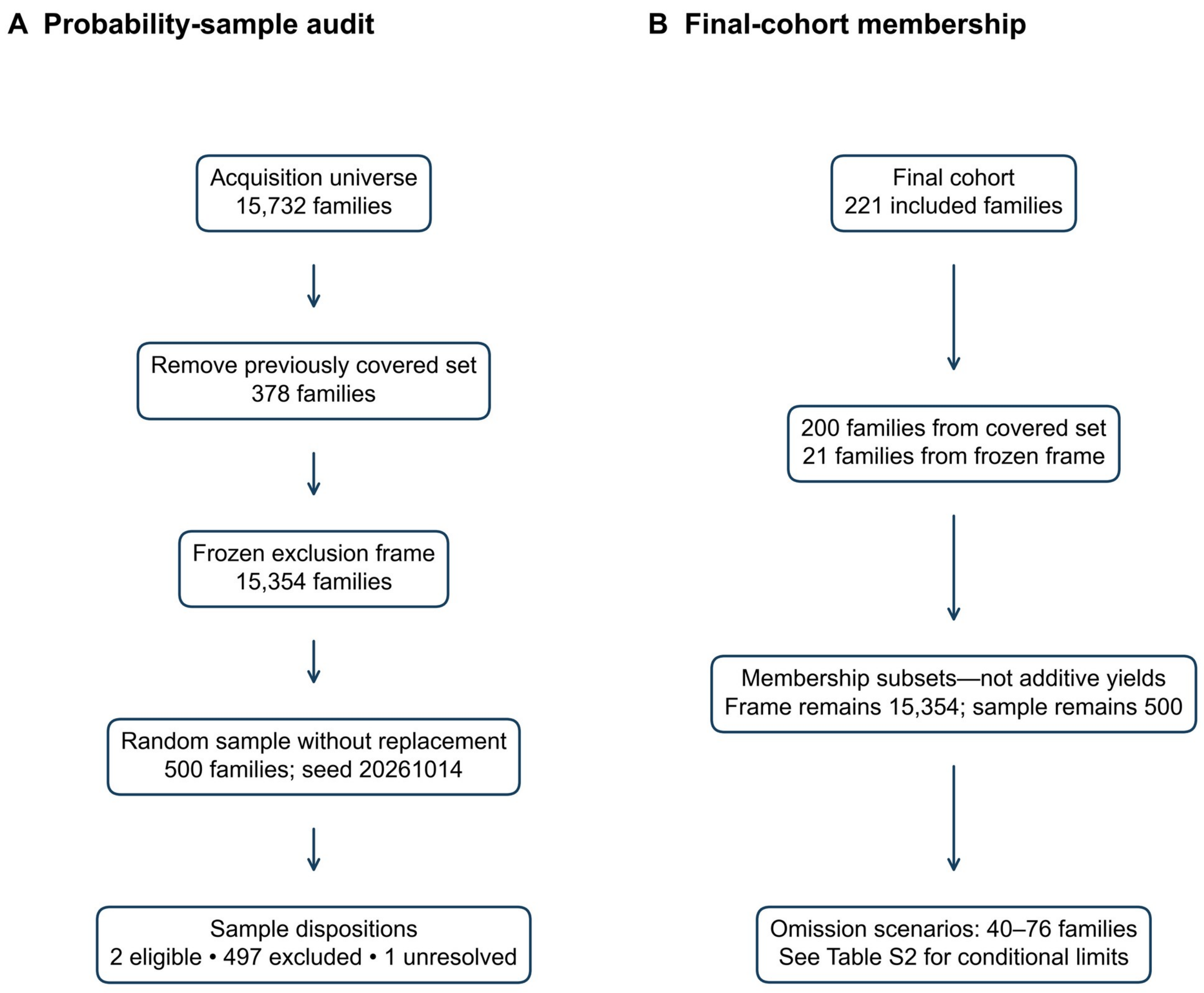

### Supplementary tables

#### Table S1: Operational variables and definitions

| Variable | Definition |
|---|---|
| Unit of analysis | Technology defined by supported mechanism and intended capability; each technology contributes once. |
| Award family | Related project records identified by family identifier; distinct from a technology and from an application. |
| Application | Exact application_id; 453 unique records linked to 209 technologies. |
| Confirmed | Funded capability/direct extension actually exercised in a living human after a qualifying same-technology award, with qualifying evidence by cutoff. |
| Not located | No qualifying evidence after documented searches and no concrete unresolved material lead. |
| Indeterminate | Specific unresolved identity, chronology, or exposure fact that could change classification. |
| Any STTR | At least one included R41 or R42 application; includes STTR-only and both-program technologies. |
| SBIR only | Included R43/R44 applications without an included STTR application. |
| Any observed Phase II | At least one included R42 or R44 application. |
| Phase I only | No included R42/R44 application; not a lifetime progression classification. |
| Award era | Minimum included fiscal year: 2005–2012 or 2013–2020. |
| Total nominal support | Sum of deduplicated qualifying application amounts; US dollars; no inflation adjustment. |
| Observation opportunity | Days from earliest valid included budget start through October 8, 2026, divided by 365.25. |
| Pre-included-start implementation | True when verified; otherwise unknown (blank/null), not false. |
| Company component | Transitive grouping through exact normalized recipient identities. |
| Funded-purpose category | One primary category per included technology; category codes appear in Table S4. |

#### Table S2: Screening omission scenarios

| Quantity | Unresolved excluded | Unresolved eligible |
|---|---|---|
| Exclusion-frame families | 15,354 | 15,354 |
| Probability-sample families | 500 | 500 |
| Sample eligible / excluded / unresolved | 2 / 497 / 1 | 2 / 497 / 1 |
| Eligible sample count used | 2 | 3 |
| Estimated eligible families in frame | 61 | 92 |
| Raw conditional exact hypergeometric limits | 8–218 | 20–264 |
| Deduplicated known eligible frame families | 21 | 24 |
| Known/scenario-assumed eligible absent families added | 0 | 8 |
| Estimated remaining omissions, families | 40 | 76 |
| Estimated family coverage | 84.5% | 74.4% |

All counts and estimates are family-level quantities. Frame totals include known recoveries. The combined transformed classification-sensitivity envelope is 0–248 remaining omitted families and 47.1%–100.0% coverage; it reflects the scenario and feasible-set constraints described in Methods S4.

#### Table S3: Ascertainment states within exploratory categorical groups

| Group | N | C | NL | I | C, % | Wilson 95% CI, % |
|---|---|---|---|---|---|---|
| any STTR | 43 | 7 | 32 | 4 | 16.3 | 8.1–30.0 |
| SBIR only | 166 | 24 | 124 | 18 | 14.5 | 9.9–20.6 |

| | | | | | | |
|---|---|---|---|---|---|---|
| any Phase II | 98 | 21 | 64 | 13 | 21.4 | 14.5–30.5 |
| Phase I only | 111 | 10 | 92 | 9 | 9.0 | 5.0–15.8 |
| 2005–2012 | 111 | 21 | 79 | 11 | 18.9 | 12.7–27.2 |
| 2013–2020 | 98 | 10 | 77 | 11 | 10.2 | 5.6–17.8 |

C=confirmed; NL=not located; I=indeterminate. Wilson intervals assume independent technologies. No-documented-confirmation counts for Fisher tests are NL+I. Differences and conventional raw/Holm p values appear in main Table 3. Funding forms the fourth test in the Holm family.

**Table S4: Included technology roster**

Rows enumerate the 209 included technologies. Stable study IDs were retained across cohort development and are nonconsecutive. C=confirmed; NL=not located; I=indeterminate. O=neuro-oncology/tumor-directed; S=spine surgery/reconstruction; V=cerebrovascular/stroke; F=functional neurosurgery/epilepsy; C=CSF/neurotrauma/neurocritical care; P=peripheral neural repair/interfaces/pain; G=implantable neural prostheses/components; K=cranial reconstruction/closure; R=neural repair/CNS-directed biologic procedures; X=cross-cutting procedural platforms.

| No. | ID | Technology | NIH families | Cat. | State |
|---|---|---|---|---|---|
| 1 | T001 | Peripheral neural interfaces and neuropathic pain | NS061603 | P | NL |
| 2 | T002 | Adjunct neuroprotection during stroke thrombectomy | NS110252 | V | NL |
| 3 | T003 | Cell-assisted spinal fusion | AR052542 | S | NL |
| 4 | T004 | CREmedical tEEG language mapping | NS105267 | F | I |
| 5 | T005 | Cortical electrode planning and follow-up | NS060203 | F | NL |
| 6 | T006 | Antimicrobial CSF shunts | NS117044 | C | NL |
| 7 | T007 | Cerebral aneurysm flow-diverter coating | NS110114 | V | NL |
| 8 | T008 | Intraoperative neurophysiological monitoring | NS061387 | X | NL |
| 9 | T009 | Percutaneous peripheral nerve stimulation for knee pain (SPR) | AG052196 | P | C |
| 10 | T010 | Neural stem-cell delivery for glioblastoma | TR001789 | O | NL |
| 11 | T011 | Deep-brain stimulation electrode arrays | NS113708 | F | NL |
| 12 | T012 | Hydrocephalus shunt pressure monitoring | NS077523 | C | NL |
| 13 | T013 | InSight stereovision | CA239 | X | I |

| | | | | | |
|---|---|---|---|---|---|
| | | brain-shift updating | 843 | | |
| 14 | T014 | Blackrock high-channel implantable auditory stimulator | DC018 757 | G | I |
| 15 | T015 | Xoran reduced-field intraoperative CBCT updating | CA107 895 | X | I |
| 16 | T016 | Epilepsy surgical planning | NS093 889 | F | NL |
| 17 | T017 | Growth-factor and stem-cell binding bone grafts | GM07 7753 | S | NL |
| 18 | T018 | Post-laminectomy adhesion prevention | AR056 493 | S | NL |
| 19 | T019 | Implant encapsulation for neural interfaces | DC018 261 | G | NL |
| 20 | T020 | DBS surgical targeting atlas (CRAVE) | MH10 0007, NS063 705 | F | C |
| 21 | T021 | Multimodal scanning-fiber endoscope | CA211 086 | O | C |
| 22 | T022 | Source Signal Imaging EMSE ECoG analysis | NS049 712 | F | I |
| 23 | T023 | Cerebral aneurysm embolization | NS059 250 | V | NL |
| 24 | T024 | Ultrasound guidance for spine instrumentation | AR055 834 | S | NL |
| 25 | T025 | Antifouling hydrocephalus shunts | NS103 704 | C | NL |
| 26 | T026 | Implantable CSF-shunt flow monitoring | NS056 628 | C | NL |
| 27 | T027 | Computer-controlled spinal fusion drilling | EB023 764 | S | NL |
| 28 | T028 | Cerebral aneurysm embolization | NS092 184 | V | NL |
| 29 | T029 | Artificial intervertebral disc replacement | AR078 140 | S | NL |
| 30 | T030 | Imaging-guided resection of brain metastases | CA141 817 | O | NL |
| 31 | T031 | AdV-tk glioma immunogene therapy | CA107 745 | O | C |
| 32 | T032 | Cortical recording and stimulation arrays | NS093 714 | F | NL |
| 33 | T033 | Drug-eluting peripheral nerve repair conduits | NS112 066 | P | NL |
| 34 | T034 | Deep-brain mapping for functional neurosurgery | NS054 346 | F | NL |
| 35 | T035 | Calvarial reconstruction | DE022 487 | K | NL |
| 36 | T036 | nView cone-beam tomosynthesis guidance | CA228 158 | S | I |
| 37 | T037 | ZetaFuse spinal-fusion implant | AR075 483 | S | C |
| 38 | T038 | Peripheral nerve regeneration scaffold | NS041 686 | P | NL |
| 39 | T039 | Transonic CSF flow sensor | NS049 680 | C | C |
| 40 | T040 | Epilepsy depth-electrode mapping | NS062 595 | F | NL |

| | | | | | |
|---|---|---|---|---|---|
| 41 | T041 | Osteoinduction for osteoporotic spinal fusion | AG063 627 | S | NL |
| 42 | T042 | BMP-mimetic bone regeneration | AR052 969 | S | NL |
| 43 | T043 | Targeted radiopharmaceutical therapy for glioblastoma | CA165 626 | O | NL |
| 44 | T044 | Dual-lumen hydrocephalus shunts | NS113 655 | C | NL |
| 45 | T045 | BMP-binding orthobiologic materials | GM09 5009 | S | NL |
| 46 | T046 | ShuntCheck / Micro-Pumper | HD065 429, NS067 770, NS067 772 | C | C |
| 47 | T047 | Peripheral nerve regeneration | NS089 341 | P | NL |
| 48 | T048 | Bone cement for orthopedic reconstruction | AR056 167 | S | NL |
| 49 | T049 | Intraoperative brain-tumor Raman imaging | CA171 740 | O | NL |
| 50 | T050 | Shunt revision catheter coagulation | NS051 924 | C | NL |
| 51 | T051 | Nanoparticle-enhanced glioma radiosurgery | CA134 074 | O | NL |
| 52 | T052 | Wearable fluorescence imaging (FLoupe) | CA243 600 | O | C |
| 53 | T053 | Nucleus pulposus replacement | AR056 892 | S | NL |
| 54 | T054 | Angler steerable microcatheter | HL137 521 | V | C |
| 55 | T055 | NeuraMedica DuraFuse | NS107 104 | S | I |
| 56 | T056 | Dural regeneration and repair | NS076 101 | X | NL |
| 57 | T057 | Precision neurosurgical laser | RR024 070 | X | NL |
| 58 | T058 | Brain infusion catheter | NS084 701 | X | NL |
| 59 | T059 | Wireless DBS powering and monitoring | NS115 226 | F | NL |
| 60 | T060 | MRI-guided neurosurgical navigation | CA168 271 | X | NL |
| 61 | T061 | NDI / SPR PNS—amputee pain | NS066 523 | P | C |
| 62 | T062 | Intraoperative cancer microscopy | CA189 320 | O | NL |
| 63 | T063 | Intraoperative vascular flow imaging | EB019 856 | V | NL |
| 64 | T064 | Electroporation ablation for glioblastoma | CA233 158 | O | NL |
| 65 | T065 | Spinal intraoperative neurophysiological monitoring | NS045 407 | S | NL |
| 66 | T066 | Cerebral aneurysm flow | NS074 | V | NL |

| | | | | | |
|---|---|---|---|---|---|
| | | diversion | 576 | | |
| 67 | T067 | Brain-tumor thermobrachytherapy | CA239815 | O | NL |
| 68 | T068 | Cerebral aneurysm liquid embolization | NS097069 | V | NL |
| 69 | T069 | Shunt maintenance and malfunction diagnosis | NS074616 | C | NL |
| 70 | T070 | Prism ALFF / NVU fMRI | CA173976 | O | I |
| 71 | T071 | Surgical adhesion prevention | GM090341 | S | NL |
| 72 | T072 | Cerebral aneurysm coiling | NS090821 | V | NL |
| 73 | T073 | Neurosurgical microrobotics | RR020247 | X | NL |
| 74 | T074 | Skull-base robotic drilling | EB009254 | K | NL |
| 75 | T075 | Craniosynostosis surgical planning | HD081712 | K | NL |
| 76 | T076 | TETRANITE cranial fixation | NS115386 | K | C |
| 77 | T077 | Vertebral augmentation cement | AR076299 | S | NL |
| 78 | T078 | CNS injury regeneration | NS048734 | R | NL |
| 79 | T079 | Cerebral aneurysm liquid embolization | NS100184 | V | NL |
| 80 | T080 | Growth-factor binding spinal fusion graft | GM097804 | S | NL |
| 81 | T081 | Processed peripheral nerve allograft | HD053159 | P | C |
| 82 | T082 | Occipital nerve stimulation for migraine | NS102302 | P | NL |
| 83 | T083 | Magnetic navigation for stroke thrombectomy | NS112035 | V | NL |
| 84 | T084 | BioTex laser ablation / vertebroplasty | CA101573 | O | I |
| 85 | T085 | Cerebral aneurysm stentgraft | NS054333 | V | NL |
| 86 | T086 | Stem-cell repair of peripheral nerve and spinal cord | AR050933, NS086217 | R | NL |
| 87 | T087 | NeurInsight TurboFIRE rsfMRI | NS090691 | O | I |
| 88 | T088 | Motion-compensated neurological CT | NS067745 | X | NL |
| 89 | T089 | Fluorescence probe for brain-tumor surgery | NS108845 | O | NL |
| 90 | T090 | ReVivo CEM-Plate / CEM-Cage | AR068819 | S | I |
| 91 | T091 | Chatten / Astro-Med S12X | NS046121 | F | I |
| 92 | T092 | CSF osmotic therapy for traumatic brain edema | NS043832 | C | NL |
| 93 | T093 | Peripheral nerve repair wrap (Nerve Tape) | NS097113 | P | C |
| 94 | T094 | Shunt malfunction pressure sensing | NS052939 | C | NL |

| | | | | | |
|---|---|---|---|---|---|
| 95 | T095 | Noninvasively adjustable scoliosis rods | AR067 008 | S | NL |
| 96 | T096 | Cortico-cancellous silicon-nitride implant | AR004 995 | S | C |
| 97 | T097 | Metastatic bone tumor ablation | CA112 852 | O | NL |
| 98 | T098 | Wireless epilepsy subdural electrodes | NS066 567 | F | NL |
| 99 | T099 | BOSS balloon guide catheter | NS103 670 | V | C |
| 100 | T100 | Intraoperative DBS target detection | NS064 640 | F | NL |
| 101 | T101 | DBS and cortical electrode coatings | NS103 714 | G | NL |
| 102 | T102 | DBS electrode coatings | NS083 183 | G | NL |
| 103 | T103 | Antimicrobial VP-shunt coatings | NS062 471 | C | NL |
| 104 | T104 | Stereotactic frame workflow for DBS | NS093 875 | F | NL |
| 105 | T105 | Antifouling CSF-shunt coatings | NS119 007 | C | NL |
| 106 | T106 | Neurovascular optical coherence tomography | NS100 163 | V | C |
| 107 | T107 | Visualase epilepsy ablation | NS077 693 | F | C |
| 108 | T108 | Intraoperative fluorescence-lifetime imaging | CA117 286 | O | C |
| 109 | T109 | Pediatric high-grade glioma virotherapy | CA250 883 | O | NL |
| 110 | T110 | Cranial dural sealant | GM13 0231 | X | NL |
| 111 | T111 | CSF-directed therapy for leptomeningeal metastases | CA206 744 | O | NL |
| 112 | T112 | Brain-tumor stimulated Raman microscopy | CA206 664 | O | NL |
| 113 | T113 | Glioma immunogene therapy | CA117 291 | O | NL |
| 114 | T115 | Robotic endoscopy for ETV/CPC | EB014 063 | C | NL |
| 115 | T116 | Speech assessment to optimize DBS | AG033 523 | F | NL |
| 116 | T117 | MRI-guided intracranial intervention | CA177 205, NS110 164 | X | NL |
| 117 | T118 | ALGEL alginate endovascular embolic material, later called Gore Embolic Liquid (GEL). | HL070 391 | V | I |
| 118 | T119 | Air Barrier System | GM09 5005 | X | C |
| 119 | T120 | CSF-leak diagnosis | NS112 064 | C | NL |
| 120 | T121 | Closed-loop hydrocephalus management | NS076 056 | C | NL |
| 121 | T122 | Cervical myelopathy | AG034 | S | NL |

| | | | | | |
|---|---|---|---|---|---|
| | | imaging and treatment planning | 732 | | |
| 122 | T123 | Needle-based tumor-bed radiotherapy | CA112 683 | O | NL |
| 123 | T124 | Piezoelectric spinal fusion implants | AR070 088 | S | NL |
| 124 | T125 | Cerebral aneurysm embolization | NS107 048 | V | NL |
| 125 | T126 | CT-based cranial and spinal surgical navigation | CA112 966 | X | NL |
| 126 | T127 | Presurgical functional mapping | NS117 226 | X | NL |
| 127 | T128 | Recurrent laryngeal nerve repair | DC017 634 | P | NL |
| 128 | T129 | Peripheral nerve regeneration conduits | NS067 771 | P | NL |
| 129 | T130 | Antimicrobial CSF shunts | NS071 776 | C | NL |
| 130 | T131 | Fluorescence-guided photodynamic tumor therapy | CA173 980, CA247 127 | O | NL |
| 131 | T132 | Neurapheresis—subarachnoid hemorrhage | NS110 247 | V | C |
| 132 | T133 | Intracranial pressure and cerebral flow monitoring | HL063 554 | C | NL |
| 133 | T134 | Tissue-engineered peripheral nerve grafts | NS108 869 | P | NL |
| 134 | T135 | Drug-eluting aneurysm coils | NS107 111 | V | NL |
| 135 | T136 | nanOss Bioactive 3D scaffold | AR052 226 | S | C |
| 136 | T137 | Cranial surgical hemostasis | NS070 331 | X | NL |
| 137 | T138 | Brain-tumor presurgical functional mapping | GM12 5438 | O | I |
| 138 | T139 | Wrong-level spine surgery prevention | AR076 830 | S | NL |
| 139 | T140 | Deep-brain stimulation arrays | NS060 269 | F | NL |
| 140 | T141 | Preoperative spinal fusion biomechanical risk assessment | AR064 613 | S | NL |
| 141 | T142 | Functional connectivity mapping for brain-tumor treatment | NS081 926 | O | NL |
| 142 | T143 | Wireless intracranial epilepsy monitoring | NS055 421 | F | NL |
| 143 | T144 | DBS electrode placement for addiction | DA051 265 | F | NL |
| 144 | T145 | Clot visualization for stroke intervention | HL095 211 | V | NL |
| 145 | T146 | Cooling adjunct for stroke thrombectomy | NS073 291, NS095 573 | V | NL |
| 146 | T147 | Kinesia / CLOVER-DBS programming | AG033 520, | F | C |

| | | | | | |
|---|---|---|---|---|---|
| | | | AG033 947 | | |
| 147 | T148 | Nanostructured spinal fixation implants | AG049 514, AR066 979 | S | I |
| 148 | T149 | CSF filtration and antifungal delivery for meningitis | AI120 304, AI120 751 | C | NL |
| 149 | T150 | WEB17 intrasaccular implant | NS071 766, NS097 038 | V | C |
| 150 | T151 | Bone-healing growth-factor gel | AR052 189 | S | NL |
| 151 | T152 | Point-of-care stem-cell enrichment for grafting | AR054 229 | S | NL |
| 152 | T153 | Hemostatic bone graft | AR056 177 | S | NL |
| 153 | T154 | Dural brachytherapy plaque | CA092 924 | O | C |
| 154 | T155 | Functional mapping near brain neoplasms | CA113 186 | O | NL |
| 155 | T156 | Nanoparticle-enhanced brain-tumor radiotherapy | CA192 702 | O | NL |
| 156 | T157 | Magnetic-particle neuromodulation for addiction | DA045 398 | F | NL |
| 157 | T158 | Postoperative spine pain measurement | DA046 985 | S | NL |
| 158 | T161 | Point-of-care adipose stem-cell enrichment | GM09 3462 | S | NL |
| 159 | T162 | Peripheral nerve-muscle interface for prosthetic control | HD049 265 | G | NL |
| 160 | T166 | Medical-device lubricity coatings | HL093 928 | X | NL |
| 161 | T167 | Microcatheter localization | HL093 935 | X | NL |
| 162 | T173 | Carotid stent for stroke prevention | HL140 895 | V | NL |
| 163 | T174 | Parkinson advanced-therapy selection and adjustment | MD01 3767 | F | NL |
| 164 | T176 | Intracerebral Microinjection Instrument | NS036 869 | X | C |
| 165 | T177 | Noninvasive intracranial pressure assessment | NS046 824 | C | NL |
| 166 | T178 | Epileptogenic-zone localization | NS051 056 | F | NL |
| 167 | T179 | Intrathoracic pressure regulator | NS054 372 | C | C |
| 168 | T180 | Bounded electrical impedance tomography | NS056 758 | X | C |
| 169 | T181 | Mimosa acoustic ICP assessment | NS058 065 | C | C |
| 170 | T182 | Epileptogenic tissue | NS060 | F | NL |

| | | | | | |
|---|---|---|---|---|---|
| | | localization | 217 | | |
| 171 | T183 | Implantable wireless subdural EEG | NS061 604 | F | NL |
| 172 | T184 | BradyXplore bradykinesia features | NS065 554 | F | C |
| 173 | T185 | Acoustic intracranial pressure monitoring | NS066 539 | C | NL |
| 174 | T186 | Simultaneous dense-array EEG/fMRI | NS071 988 | X | NL |
| 175 | T187 | Continuous CSF-shunt flow monitoring | NS074 486 | C | NL |
| 176 | T188 | Traumatic brain-injury intracranial pressure prediction | NS076 107 | C | NL |
| 177 | T189 | Subdermal ambulatory EEG | NS076 270 | G | NL |
| 178 | T190 | DBS programming optimization | NS081 902 | F | I |
| 179 | T191 | Focal cortical dysplasia detection | NS083 101 | F | NL |
| 180 | T192 | Post-cardiac-arrest neuroprotection | NS089 196 | C | NL |
| 181 | T193 | Cerebral aneurysm coil coatings | NS095 455 | V | NL |
| 182 | T194 | Intracranial trauma monitoring | NS100 313 | C | NL |
| 183 | T196 | Neural electrode insertion | NS105 500 | X | NL |
| 184 | T197 | Intra-arterial stroke intervention | NS107 108 | V | NL |
| 185 | T198 | Locoregional pain neural ablation | NS110 272 | P | NL |
| 186 | T199 | Aneurysm recurrence surveillance | NS115 253 | V | NL |
| 187 | T200 | Intracranial aneurysm detection and rupture-risk assessment | NS115 314 | V | NL |
| 188 | T201 | Intercostal nerve block for thoracotomy pain | NS115 460 | P | NL |
| 189 | T202 | Neurofibromatosis type 2 tumor bacteriotherapy | NS120 338 | O | NL |
| 190 | T203 | Cellarium CNS-specific implantable glioma vaccine | NS071 879 | O | I |
| 191 | T204 | CNS stem-cell transplantation (HuCNS-SC) | AG022 736 | R | C |
| 192 | T205 | Characterizing marrow derived cells for neural therapy | AG026 168 | R | NL |
| 193 | T206 | hD2b-engineered human neural stem-cell intracranial transplantation | AG056 186 | R | NL |
| 194 | T207 | Somatostatin (SST)-GABAergic Interneuron Therapy for Alzheimer's Disease with ApoE4 | AG060 873 | R | NL |
| 195 | T208 | Actuated Medical BIFOR / OsteoAccess | CA139 774 | O | NL |

| | | | | | |
|---|---|---|---|---|---|
| | | low-force bone biopsy | | | |
| 196 | T209 | Small-molecule conversion of fibroblasts to neurons (hCiN) | DA041 997 | R | NL |
| 197 | T210 | Aculight optical cochlear nerve stimulator and compact diode-driver platform | DC008 515 | G | I |
| 198 | T211 | Front Edge NanoEnergy fully implantable thin-film rechargeable lithium battery | DC009 110 | G | NL |
| 199 | T212 | Vixar 1860 nm VCSEL sources for implanted optical neuroprostheses | DC012 776 | G | NL |
| 200 | T213 | iotaMotion implanted robotic cochlear electrode positioner (IOTA-Progress) | DC016 246 | G | I |
| 201 | T214 | CNS AAV-IDUA gene delivery (RGX-111 lineage) | DK094 538 | R | C |
| 202 | T216 | Acoustic MedSystems multiplexed-DOF image-guided needle robot | GM10 0589 | X | NL |
| 203 | T217 | NDI battery-powered wireless upper-limb motor neuroprosthesis | HD049 960, NS055 423 | G | I |
| 204 | T220 | Optogenetic control over transgene expression for the therapy of brain and spine | MH11 0273 | R | NL |
| 205 | T222 | Stem Cells for Neonatal Hypoxic-Ischemic Brain Injury | NS055 606 | R | I |
| 206 | T223 | Synapse translumenal NOTES phrenic motor-point mapping and electrode-delivery instrumentation | NS057 845 | P | NL |
| 207 | T224 | Astrocyte-restricted precursors for cell therapy and drug screening | NS067 719 | R | I |
| 208 | T225 | Ferro wireless optical DBS / optogenetic neuromodulation | NS070 453 | F | NL |
| 209 | T226 | RXi/Thera SOD1-targeting self-delivering rxRNA CNS therapy | NS074 671 | R | NL |

**Table S5: Confirmed implementation evidence and source links**

| **Technology and qualifying setting** | **Capability, exposure, and chronology grounds** |
|---|---|
| T009<br>Percutaneous peripheral nerve stimulation for knee pain (SPR)<br>(Neurosurgical scope) | Seven participants enrolled March–October 2016 received percutaneous SPR femoral/sciatic leads and delivered stimulation, including during rehabilitation. The primary report dates exposure after the August 2015 award and demonstrates the funded postoperative pain interface, not simply |

| | |
|---|---|
| | enrollment. Source 1 |
| T020<br>DBS surgical targeting atlas (CRAVE)<br>(Neurosurgical scope) | The 2011–2021 DBS series states that CRAVE atlas heat maps guided surgical targets and trajectories and final electrode positioning. This is actual procedural guidance after the September 2009 award, despite retrospective outcome analysis. Source 1 |
| T021<br>Multimodal scanning-fiber endoscope<br>(Other clinical) | The primary report documents real-time multimodal fluorescence/reflectance endoscopic imaging in 22 Barrett participants and links them to NCT03589443, actually conducted May 2018–February 2019 after the May 2017 award. Modular multiwavelength scanning and concurrent fluorescence/reflectance imaging support a direct procedural extension; systemic peptide exposure alone is not the confirming event. Source 1 Source 2 |
| T031<br>AdV-tk glioma immunogene therapy<br>(Neurosurgical scope) | Primary multicenter results document local AdV-tk administration at glioma surgery during 2006–2010, with 48 completing the regimen. The local surgical delivery implements the funded capability after the June 2004 qualifying award; oral prodrug exposure alone would not. Source 1 |
| T037<br>ZetaFuse spinal-fusion implant<br>(Neurosurgical scope) | The case report identifies ZetaFuse's small-molecule/collagen/calcium-phosphate formulation packed into cages during ACDF. Cervical stenosis diagnosed in 2022 followed by failed conservative care and surgery establishes post–September 2019 implementation by the 2024 report. Cervical use directly extends the funded lumbar fusion capability. Source 1 |
| T039<br>Transonic CSF flow sensor<br>(Neurosurgical scope) | The primary report describes a Transonic transit-time sensor connected in-line to an EVD and measuring CSF in living patients. Its five Wisconsin pediatric cases are linked to NCT00652197 and the March 2009–May 2014 interval, after September 2005. Extracorporeal shunt/EVD sensing is explicitly funded, not generic physiologic observation. The registry interval dates the pediatric subset, not necessarily all 16 patients. Source 1 Source 2 |
| T046<br>ShuntCheck / Micro-Pumper<br>(Neurosurgical scope) | The multicenter primary report dates actual thermal shunt-flow testing to April 2013–March 2016 and describes micropumper application over the implanted valve. This exercises funded shunt-patency diagnostics after March 2010 even with investigators blinded to test results. The report distinguishes 406 enrolled participants from 391 valid tests. Source 1 |
| T052<br>Wearable fluorescence imaging (FLoupe)<br>(Neurosurgical scope) | Two FLoupe prototypes imaged tumors in 11 living patients during glioma operations. The paper connects engineering, calibration and clinical applications to its funded Phase-I development sequence after August 2019; exact surgery dates remain unreported. In-situ fluorescence imaging exercises the funded capability even though standard microscopes guided resection and FLoupe was evaluated comparatively. Source 1 |
| T054<br>Angler steerable microcatheter<br>(Other clinical) | Treating interventionalists report Angler active-deflection microcatheter use in 40 peripheral intervention cases during mid-2022–mid-2023, after January 2018. Named product and coaxial mechanism connect the actual procedural navigation to the funded platform. This is qualifying cross-specialty use, not evidence of neurovascular clinical deployment. Source 1 |
| T061<br>NDI / SPR PNS—amputee pain<br>(Neurosurgical scope) | The primary randomized trial documents implanted percutaneous leads and active SPRINT stimulation for post-amputation pain during March 2015–March 2018, after September 2009. This executes the Phase-I percutaneous pain capability; the later fully implanted pulse-generator goal need not be assumed achieved. Source 1 |
| T076<br>TETRANITE cranial fixation<br>(Neurosurgical scope) | The 2026 primary conference case report describes adhesive fixation of a cranial flap in a 44-year-old patient and explicitly links the case to NCT06780852. That TETRANITE protocol began January 15, 2025, after the May 2020 award. Actual flap fixation exercises the funded delivery/fixation capability; the registry alone would not prove exposure. Source 1 Source 2 |
| T081<br>Processed peripheral nerve allograft<br>(Neurosurgical scope) | Primary results document processed Avance nerve allograft repairs in 2008–2015 after April 2007. Decellularization, CSPG removal and retained basal-lamina scaffold support the funded processed-graft identity. Actual nerve reconstruction qualifies even though the treating service is plastic/orthopedic hand surgery. Source 1 |
| T093<br>Peripheral nerve repair wrap (Nerve Tape)<br>(Neurosurgical scope) | The primary case-series abstract reports 19 people undergoing 21 Nerve Tape lingual/inferior-alveolar repairs in 2023–2025, after April 2016. Named product and microhook coaptation development support the funded |

| | |
|---|---|
| | repair capability. Oral/maxillofacial service does not invalidate actual neural repair. Source 1 |
| T096<br>Cortico-cancellous silicon-nitride implant<br>(Neurosurgical scope) | CASCADE results report 52 recipients of dense-shell/microporous-core silicon-nitride spacers among 100 analyzed participants in December 2011–October 2013. Implantation at ACDF directly extends the funded cortico-cancellous fusion architecture after April 2005; ordinary hollow silicon-nitride cages alone would not prove that architecture. Source 1 Source 2 |
| T099<br>BOSS balloon guide catheter<br>(Neurosurgical scope) | FAST-BGC reports 50 treated stroke patients, distinct from 51 consented, with BOSS support/flow arrest during mechanical thrombectomy. The paper links NCT04416659, whose actual 2021 study dates follow September 2018. Actual device-assisted revascularization, not clearance or recruitment, confirms the capability. Source 1 Source 2 |
| T106<br>Neurovascular optical coherence tomography<br>(Neurosurgical scope) | The primary report documents microcatheter-delivered nOCT acquisitions in intracranial arteries of 32 living patients, including peri-intervention device-vessel imaging. Its linked trial route began 2023-02-13, after the 2017-04-01 award; this dates at least the trial subset, not all special-access recipients. Actual intravascular imaging exercises the funded capability. Source 1 Source 2 |
| T107<br>Visualase epilepsy ablation<br>(Neurosurgical scope) | Posted NCT02844465 results identify 114 recipients of Visualase amygdala/hippocampal ablation during the 2016-12-01 to 2023-12-11 study interval, after the 2012-06-01 award. This is actual living-human epilepsy ablation, not clearance or intended enrollment. Source 1 |
| T108<br>Intraoperative fluorescence-lifetime imaging<br>(Neurosurgical scope) | The primary report describes a sterilized FLIm probe handed to the surgeon for in-vivo scanning along brain-tumor resection paths in three craniotomy patients. The documented extension from TR-LIFS and operation under the 2014 ANSI safety design bound use after the 2006 award. The human implementation was in-vivo scanning during craniotomy; exact surgical dates were unavailable. Source 1 |
| T119<br>Air Barrier System<br>(Mixed surgical) | Posted ABS trial outcomes and the clinical report document use of the local clean-air barrier over open incisions in actual implant operations after the 2010 award. This implements the funded surgical-field capability; mixed orthopedic, spinal and vascular procedures are permissible rather than requiring every recipient to be neurosurgical. Source 1 Source 2 |
| T132<br>Neurapheresis—subarachnoid hemorrhage<br>(Neurosurgical scope) | PILLAR-XT reports automated catheter-based CSF filtration in 27 treated patients. The linked actual January 2021 immediate-catheter-removal primary completion and maximum 72-hour catheter dwell support at least one use after the August 2019 award. The evidence does not date all 27 procedures or establish an exact first-use date. Source 1 Source 2 |
| T136<br>nanOss Bioactive 3D scaffold<br>(Neurosurgical scope) | Posted NCT01829997 results document 22 started recipients of nanOss Bioactive 3D in lumbar fusion in the April 2013-April 2018 study window, after the September 2005 award. The E-Matrix/FortrOss/NB3D material-and-naming chain supports a direct scaffold extension; neither sponsor ownership nor FDA predicate status alone is the identity basis. Source 1 |
| T147<br>Kinesia / CLOVER-DBS programming<br>(Neurosurgical scope) | CLOVER-DBS documents iterative Kinesia sensor feedback used to choose and apply stimulation settings in 23 DBS recipients. The paper identifies NCT03037398, whose January 2017 start is after the September 2008 award. This is actual programming capability; the registry's larger enrollment must not be substituted for the published cohort. Source 1 Source 2 |
| T150<br>WEB17 intrasaccular implant<br>(Neurosurgical scope) | The primary WEB17 report documents 40 patients/46 aneurysms treated December 2016-September 2017, after the June 2016 award for WEB-017 microcatheter-compatible development. Actual intrasaccular deployment exercises that configuration-specific capability. Source 1 |
| T154<br>Dural brachytherapy plaque<br>(Neurosurgical scope) | The primary report describes eight intraoperative dural treatments during February 2000–February 2002, progressing from iridium to liquid yttrium and finally four yttrium-foil uses. The chronology assessment places at least one final foil use after the separately verified September 2001 R43 award. Exact post-award recipient count is unknown; the 2005 Phase II award does not date these earlier procedures. The journal report was published in 2003. NIH award Primary report |
| T176<br>Intracerebral Microinjection Instrument | The primary stroke transplantation report identifies the FHC side-exit Intracerebral Microinjection Instrument in cohort C, three recipients |

| | |
|---|---|
| (Neurosurgical scope) | receiving 45 deposits across three cannula tracks. Its 2013-2016 treatment sequence follows the observed 2008-2009 awards and directly exercises spatial intracerebral delivery, not merely use of related cells. Source 1 |
| T179<br>Intrathoracic pressure regulator<br>(Neurosurgical scope) | The primary clinical extract documents insertion into ventilator circuits and 20 applications in a trial commencing July 2009, after the February 2006 award. Active ICP-lowering treatment implements the funded capability. Twenty is an application count, not a unique-patient count. Source 1 |
| T180<br>Bounded electrical impedance tomography<br>(Nonclinical human) | Active bEIT head-current measurements were performed in four adults. Phase II NIH aims place the human conductivity measurements in Phase I after the September 2006 start. This nonclinical human work exercises the funded head-conductivity measurement capability. Exact session dates and healthy status are not established. Conference paper Institutional publication record |
| T181<br>Mimosa acoustic ICP assessment<br>(Nonclinical human) | The primary report describes Mimosa HearID DPOAE/middle-ear measurements made January-June 2008, after the April 2007 award. Healthy-participant tilt-table testing exercised the funded experimental method for eliciting ICP-related acoustic changes, establishing nonclinical human use of the acoustic measurement capability. Source 1 |
| T184<br>BradyXplore bradykinesia features<br>(Neurosurgical scope) | NCT02358876 directly identifies BradyXplore/5R44NS065554 and reports 32 completed clinic assessments (30 completed home use), February-July 2015 after the 2009 award. Its clinic protocol includes DBS on/off/washout and reactivation with synchronized motion assessment. This exercises the funded DBS-response evaluation capability; home monitoring or earlier medication-only studies alone would not suffice. Source 1 |
| T204<br>CNS stem-cell transplantation (HuCNS-SC)<br>(Neurosurgical scope) | Selden 2013 primary abstract documents six children receiving HuCNS-SC grafts into cerebral hemispheres/lateral ventricles. Taupin 2006 explicitly connects the proprietary HuCNS-SC therapy across NCL and AD and dates trial initiation after March 2006 IND approval, later than the September 2005 award. The funded capability was intracranial grafting of unmodified purified CNS stem cells. The cross-indication linkage concerns this graft capability. Donor-line differences described by Marsh 2017 restrict transfer of efficacy and safety findings across products. Source 1 |
| T214<br>CNS AAV-IDUA gene delivery (RGX-111 lineage)<br>(Neurosurgical scope) | Wang et al. 2026 primary clinical report documents CT-guided cisterna-magna needle access and actual RGX-111 injection in a child with MPS I; it carries the exposure claim. Grant-linked AAV9-IDUA route work and the REGENXBIO lineage support a direct CNS-delivery extension of the award. Contemporaneous 2019/2020 sponsor reports corroborate the single-patient program and post-2012 chronology; publication year and registry start do not independently date treatment. The November 2019 announcement is an upper bound, not an exact dosing date. Source 1 |

Evidence for the 31 confirmed technologies, including capability linkage, human exposure, chronology, and supporting source links. Participant counts refer to the individual reports. T076 is supported by a primary conference case report.

**Table S6: Indeterminate implementation evidence**

All 22 technologies remain included in the primary denominator. These are concrete unresolved endpoint facts, rather than verified nonuse. Shared company identity, common mechanism ancestry, or later publication alone cannot settle the missing linkage or chronology. Source identifiers and links are retained with the archived analytical records.

| Technology | Unresolved fact and evidentiary grounds |
|---|---|
| T004 | A 2022 report evaluates language lateralization, but acquisition after the |

| | |
|---|---|
| CREmedical tEEG language mapping | May 2019 award is not established. A separate 2026 speech classifier records human signals without demonstrating the funded localization capability. Missing fact: a post-award implementation interval and actual execution of the funded mapping function. |
| T013<br>InSight stereovision brain-shift updating | The clinical report describes stereovision brain-shift imaging, but the interval after the qualifying award and contemporaneous execution of the funded image-updating functions remain unresolved. Missing fact: a dated qualifying implementation interval and configuration-level functional evidence. |
| T014<br>Blackrock high-channel implantable auditory stimulator | Human auditory-nerve stimulation after the June 2020 award is supported, but sources identify an initial 12-channel MED-EL-based system. Missing fact: linkage of the configuration actually used to the separately funded 60/128-channel stimulator, enclosure, and feedthrough capability or a supported direct extension. |
| T015<br>Xoran reduced-field intraoperative CBCT updating | Living-human xCAT procedures during July 2008–September 2009 follow the award. Missing fact: evidence that the clinical system executed the funded reduced-field regional updating capability, or a supported direct extension, rather than generic cone-beam CT imaging. |
| T022<br>Source Signal Imaging EMSE ECoG analysis | A human EMSE planning episode is a concrete lead, but its date and the software functions used are unresolved. Missing fact: a post-2004 episode implementing the funded ECoG localization/visualization capability, distinguished from EMSE spectral analysis and separate Persyst rendering. |
| T036<br>nView cone-beam tomosynthesis guidance | A primary AAPM report describes clinical nView guidance but does not date the cohort after September 2017. A December 2020 hospital first-use announcement does not independently date those Stanford cases. Missing fact: dated primary clinical documentation establishing a qualifying post-award interval. |
| T055<br>NeuraMedica DuraFuse | A February 2026 surgeon account describes DuraFuse patient closure; cadaver and animal reports do not establish living-human implementation. Missing fact: a primary clinical report or posted results documenting actual closure in a living recipient after June 2018 and publicly available by the cutoff. |
| T070<br>Prism ALFF / NVU fMRI | Twelve ALFF/NVU cases were selected from a 2012–2016 population straddling the July 2013 award. Corrected maps alone do not establish contemporaneous implementation. Missing fact: post-award patient dates and execution of correction during procedural evaluation, rather than retrospective image reanalysis. |
| T084<br>BioTex laser ablation / vertebroplasty | NCT00688038 has post-award actual study dates, but completed status, estimated enrollment, and a planned laser protocol do not establish treatment. Missing fact: primary or posted results identifying actual recipients, dates, and the funded MR-guided ablation/stabilization capability or a documented extension. |
| T087<br>NeurInsight TurboFIRE rsfMRI | The five-patient TurboFIRE report describes real-time processing that changed ongoing scans, supporting actual human capability use. Preliminary patient work preceded the December 2015 award. Missing fact: a dated acquisition interval or supported development sequence establishing post-award implementation. |
| T090<br>ReVivo CEM-Plate / CEM-Cage | CEM-Plate/CEM-Cage implantation claims and a 2022 trial start are material leads. Missing fact: primary clinical reporting or posted results available by the cutoff that corroborate actual post-award implantation of the funded configuration. Anticipated 2027 results do not satisfy the cutoff. |
| T091<br>Chatten / Astro-Med S12X | A 2007–2011 Grass/Astro-Med series documents cortical stimulation after the May 2003 award. Missing fact: configuration-level linkage to the funded Chatten switching and rapid post-stimulation EEG-recovery capability, rather than generic stimulation and recording. |
| T118<br>ALGEL alginate endovascular embolic material, later called Gore Embolic Liquid (GEL). | A 2006 trade report identifies a treated MAGELAN patient and states that GEL was formerly ALGEL. Missing fact: qualifying primary clinical reporting or posted results corroborating actual ALGEL/GEL administration after funding and by the cutoff. The lineage/treatment lead remains unresolved. |
| T138<br>Brain-tumor presurgical functional mapping | The MLP report uses 2014/2015 scans preceding the September 2017 award. Later publication or reprocessing does not date living-patient use. |

| | |
|---|---|
| | Missing fact: a post-award participant episode implementing the funded Cirrus/MLP mapping capability during clinical evaluation. |
| T148<br>Nanostructured spinal fixation implants | A surgeon-authored Nanovis case report describes L3–L5 TLIF/pedicle-screw implantation and one-year outcome. Missing fact: calendar implantation chronology, verified pre-cutoff public availability, and qualifying primary-report provenance for the Nano FortiCore/FortiFix case. Clearance dates and URL directories do not supply procedure dates. |
| T190<br>DBS programming optimization | The Phase II abstract describes Phase I patients undergoing algorithm-guided DBS monopolar surveys with changes to stimulation settings. Missing fact: a qualifying primary report or posted results establishing post-award exposure chronology. This procedural-use lead remains indeterminate. |
| T203<br>Cellarium CNS-specific implantable glioma vaccine | A primary-supported melanoma WDVAX implementation lead exists, but shared scaffold ancestry does not establish the funded CNS-specific Cellarium capability. Missing fact: a documented formulation, manufacturing, development, or grant-outcome bridge supporting WDVAX as the same capability or a direct extension. No qualifying intracranial Cellarium report was located. |
| T210<br>Aculight optical cochlear nerve stimulator and compact diode-driver platform | A 2009 primary conference abstract reports living-human auditory-nerve optical stimulation, but device lineage and treatment timing relative to September 2006 are unresolved. Other optical-stimulation systems do not establish Aculight identity. Missing fact: methods and chronology linking the human exposure to the funded OCS/direct extension, or informative qualifying results from NCT05110183 identifying that system. |
| T213<br>iotaMotion implanted robotic cochlear electrode positioner (IOTA-Progress) | Actual post-award iotaSOFT use is established, but its temporary intraoperative insertion drive differs from the retained postoperative IOTA-Progress positioner. Missing fact: a supported development/capability bridge from DC016246 to the clinically used drive, or a qualifying human IOTA-Progress report after April 2017. Company association alone is insufficient. |
| T217<br>NDI battery-powered wireless upper-limb motor neuroprosthesis | Living-human MICROPULSE implantation is established, but the relationship between the funded upper-limb motor device and the shoulder-pain system remains unresolved. Missing fact: design, engineering-history, or award-linked documentation connecting NS055423/HD049960 to the implanted model. A different indication could qualify if that capability bridge were supported. |
| T222<br>Stem Cells for Neonatal Hypoxic-Ischemic Brain Injury | Systemic IV MultiStem use does not exercise the funded neural delivery capability. Penn's 2012 report describes coronary adventitial catheter micro-injection in 19 recipients enrolled September 2008–February 2010. Missing fact: a documented direct-extension bridge from the NS055606-funded neural procedural capability to that delivery system, or qualifying neural-delivery human evidence. |
| T224<br>Astrocyte-restricted precursors for cell therapy | A 2025 investigator conference report describes Q-Cells GRP spinal transplantation beginning in late 2022. The award develops astrocyte-restricted precursors, whereas Q-Cells generate astrocytes and oligodendrocytes. Missing fact: a supported identity/direct-extension bridge from the funded ARP capability to the clinical GRP product. Shared recipient identity is insufficient. |

**Table S7: Unresolved eligibility families outside the outcome denominator**

These eight families are not the 22 included technologies with indeterminate implementation. No implementation state is assigned while eligibility is held.

| Family | Unresolved funded-purpose or route evidence |
|---|---|
| MH119979 | Both funded records describe a programmable supercontinuum source, two unnamed prototype systems with and without neural intervention, and possible operating-room uses. The grant-acknowledged Liu report demonstrates optical excitation/imaging in harvested mouse hippocampal slices. Its stereotaxic injection prepares specimens; the laser is not shown performing or guiding that surgery. The relationship between the funded |

| | |
|---|---|
| | source and a qualifying neural intervention remains unresolved. |
| NS103596 | The earlier abstract permits NPEX delivery intravenously or directly into CSF, whereas later IND-enabling aims describe a broadly administered neurorestorative biologic without a delivery route. Neural-cell ancestry and adjunctive use after thrombectomy alone are insufficient; whether the funded work actually develops a direct CSF procedural capability remains unresolved. |
| CA174280 | The funded MRI-gradient insert targets deep brain/skull-base tumors, a genuine potential cranial-oncology purpose. However the grant abstract says BBB bypass 'traumatically' while its narrative says 'without trauma'. The grant-linked focusing paper is in vitro; the related patent supplies an intranasal embodiment without proving that embodiment was the funded route. External particle propulsion alone does not resolve whether a qualifying cranial procedure is funded. The funded procedural route remains unresolved. |
| MD014092 | The grant funds an external magnetic workstation and particles to improve neuroprotectant delivery in acute ischemic stroke during transfer. Its patent AIS example specifies intravascular administration but does not distinguish peripheral IV from local/intra-arterial access; separate hepatic TAE/TACE catheter embodiments are not proof of the AIS award's route. Neurovascular disease and external magnetic propulsion are insufficient by themselves. A dedicated procedural adjunct/route remains unresolved. |
| NS115249 | The three exact NIH records fund OS2966 efficacy in porcine TBI without specifying the TBI administration route. They separately describe IV nonhuman-primate safety and an intracerebral recurrent-glioblastoma clinical program. Neither observation establishes the funded TBI route; a shared antibody is not proof of a shared procedural capability or direct extension. The TBI delivery route remains unresolved; eligibility and implementation are not inferred from the separate glioblastoma program. |
| NS113704 | The funded dIEE AAV vectors target intracellular/extracellular alpha-synuclein in Parkinson disease and are tested in mouse brains. CNS expression and bypassing the antibody BBB problem do not establish intracranial/intrathecal rather than systemic AAV administration. The direct neural-delivery possibility is material but unresolved. No clinical endpoint is inferred from vector ancestry or neurological indication. |
| NS055405 | The source states neural cell-therapy intent, but the funded Phase I application develops in-vitro expansion, characterization and banking. Engraftment and efficacy in spinal cord injury animal models are explicitly proposed for a future Phase II study, not the funded Phase I. The abstract does not establish an intended intracranial or intraspinal administration route; cell origin and disease target do not establish route. No clinical endpoint is assigned while eligibility remains unresolved. |
| NS079001 | The funded Phase I abstract explicitly includes efficacy after transplant into spinal cord injury animal models, with neural induction and therapeutic intent. It does not specify an intracranial or intraspinal administration route. The SCI disease model alone cannot establish that route; expansion to ALS and Parkinson's models is a future Phase II proposal. No clinical endpoint is assigned while eligibility remains unresolved. |

Source-access records for two held families: NS055405; NS079001.

#### Table S8: Indeterminate classification scenario

| Scenario | Confirmed technologies | Proportion, % |
|---|---|---|
| Observed classifications | 31/209 | 14.8 |
| All 22 indeterminate technologies become confirmed | 53/209 | 25.4 |

The hypothetical scenario retains all 209 technologies and leaves the 156 not-located classifications unchanged. It isolates the effect of resolving the 22 indeterminate cases as confirmed. It does not provide an upper bound on actual implementation among all technologies or correct for screening omissions.
